# STARSHIP: Study of Telomeres And Role of Sex Hormones In Pulmonary fibrosis

**DOI:** 10.64898/2026.08.03.26359301

**Authors:** Anna Duckworth, Julia K. Prague, Bea Knight, Kevin Norris, Holly Emms, Sophie Goodrum, Charlotte S. Crook, Ross Sayers, Matt Steward, Hannah Thould, Andrew Savill, Jessica Mandizha, Sarah Lines, Anna Barnes, John Kirkwood, Howard Almond, Katie Lunnon, Mark A. Lindsay, Jess Tyrrell, Stefan Stanel, Duncan M. Baird, Anne-Marie Russell, Pilar Rivera-Ortega, Michael A. Gibbons, Chris J. Scotton

## Abstract

**Background:** Fibrotic interstitial lung disease (F-ILD) has high mortality. Evidence suggests short telomere causality and sex hormone interactions. STARSHIP aimed to assess feasibility for future F-ILD sex hormone trials.

**Methods:** Leukocyte telomere length (LTL), complete blood count, sex hormone (testosterone and oestrogen), sex hormone binding globulin (SHBG) and albumin concentrations were determined in 102 F-ILD outpatients (age 49-89, male N=80 [78%]) and age/sex-matched controls (ASMCs). Patients undertook routine pulmonary function tests, 93 (91%) participated in bespoke telephone interviews. Survival was assessed at median 33 (28-39) months.

**Results:** 77/79 (97.4%) male patients had haemoglobin and haematocrit below the upper reference limit. Mean LTL was shorter for patients than ASMCs (4.57kb [95%CI:4.46-4.69] vs 4.78kb [95%CI:4.67-4.89]; p<0.006). SHBG concentrations were higher for patients. Mean bioavailable testosterone was lower for N=80 male patients than ASMCs (4.95nmol/L [95%CI:4.50-5.41] vs 6.40nmol/L [95%CI:5.82-6.98]; p<0.0001). Post-menopausal oestrogen concentrations were low for female patients and controls. Mean free androgen index (FAI) was low for female patients but not ASMCs (mean 0.51 [95%CI:0.35-0.67] vs 1.23 [95%CI:0.77-1.69]; p=0.0036, N=22). Age/BMI-adjusted bioavailable testosterone concentration in male patients correlated with both DLCO% (*β*=3.31, p=2.4×10^-4^) and FVC% (*β*=2.76, p=0.0030). FVC% associated with FAI in females (*β*=34.3, p=0.0029). In all-confounder-adjusted Cox analysis, low free testosterone associated with mortality (HR=2.66, p=0.023, N=77) in male patients. Lower FAI (adjusted for age/lung function) suggested similar effects but more studies needed for females (HR=3.59, p=0.22, N=18).

**Conclusions:** ILD patients have low sex hormones concentration(s), which associated with reduced lung function and survival. Sex hormone supplementation studies are needed.

**Key messages:** Using population data, we have previously shown a three-way association between short telomeres, low sex hormones and higher pulmonary fibrosis (PF) prevalence. However, no clinical data on sex hormone concentrations for PF patients and links with lung function and survival have been reported. Here we show, in a prospective clinical study, that male and female patients presenting to the interstitial lung disease clinic with PF have lower sex hormone concentrations than age- and sex-matched controls, which is associated with worse lung function and reduced survival. The findings highlight the need to explore potential benefits of hormone supplementation in PF.

## INTRODUCTION

The key pathology in interstitial lung disease (ILD) is inflammation and/or fibrosis^1^. Fibrotic ILD has 3.0 years median survival^2^. No current and upcoming antifibrotic treatments for idiopathic pulmonary fibrosis (IPF)^3,4^ and progressive fibrosing ILDs (PF-ILD)^5^ arrest lung function decline or reverse progression, and all have side-effects.

Short telomere causality in both familial^6,7^ and idiopathic/sporadic^8^ pulmonary fibrosis is well-established. Association of short telomere length with worse survival across subtypes^9–11^ informed our rationale to include all patients with fibrotic ILD. Here ‘pulmonary fibrosis’ describes all ILD subtypes with a fibrotic pattern.

Using UK Biobank data, we previously reported multiple consistent associations between short telomeres, sex hormones and pulmonary fibrosis^12^. The literature supports a direct mechanistic role for sex hormones in telomere maintenance^13^.

Importantly, sex hormone binding globulin (SHBG) binds androgens and oestrogens in blood and regulates their access to target tissues^14^. In males, approximately 44% of total testosterone is tightly bound to SHBG, with 54% loosely bound to albumin and the remaining 0.5-3% unbound or ‘free’. The latter two fractions constitute ‘bioavailable testosterone’; reports vary about differential impact on cell activity, and it may be tissue-dependent. The proportions of free testosterone and oestrogen in blood are inversely related to SHBG concentration^15^. Measurements of serum SHBG, albumin and total testosterone are used to calculate free testosterone concentration in males^16^.

In females, oestrogen is also bound by SHBG but clinically no adjustment is made, despite evidence of abnormally low oestrogen exposures in women with high SHBG^17^. A low ‘free oestradiol index’ (oestradiol/SHBG)^18^ can be used. Similarly, we judged free androgen index (derived in pre-menopausal women) to be the appropriate surrogate marker for post-menopausal bioavailable testosterone.

Testosterone levels can regulate erythrocytosis; supra-physiological dosing of exogenous testosterone can cause polycythaemia and/or hyper-viscosity. Lung disease can also cause secondary polycythaemia due to hypoxia. Measurement of haemoglobin concentration and haematocrit (red cell to total blood volume ratio) in a full blood count enables automated monitoring of haematological indices from a single venepuncture.

STARSHIP (Study of Telomeres And Role of Sex Hormones In Pulmonary fibrosis) set out to assess red blood cell safety margins for erythrocytosis as primary analyses and telomere length and sex hormone parameters as secondary analyses. These findings are needed to gauge the potential for future therapeutic trials of sex hormone supplementation in fibrotic ILD.

## METHODS

### Study design and participants

STARSHIP was a pragmatic case-control study designed to include all qualifying and consenting patients attending outpatient clinic within the study period: 102 patients (Male:Female ratio 80:22, age-range male 49-86y and female 64-89y) with stored serum samples from 102 age- and sex-matched controls (ASMCs). Inclusion criteria required a multi-disciplinary team diagnosis of IPF or a fibrotic ILD (F-ILD), described here as ‘pulmonary fibrosis’ (PF), and routine clinic appointment attendance at our regional ILD centre (Exeter, UK) with same day pulmonary function tests (**Figure 1**).

**Figure 1:**
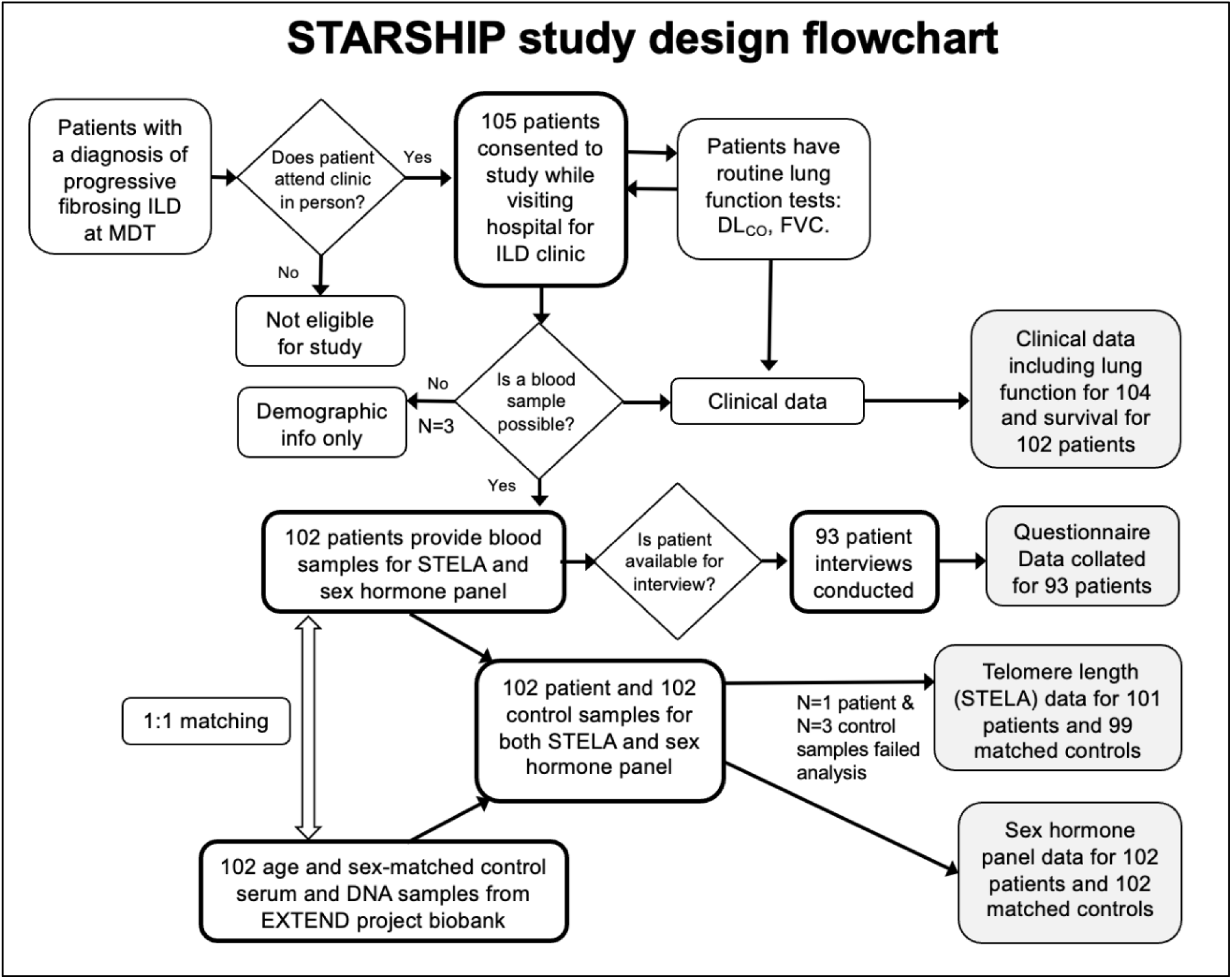
STARSHIP study design showing patient and sample numbers at each stage, with key results datasets shaded. Abbreviations: MDT – multi-disciplinary team, ILD – interstitial lung disease, DLCO – diffusing capacity for carbon monoxide, FVC – forced vital capacity, STELA – single telomere length analysis, EXTEND – Exeter ten thousand. 105 patients with PF were recruited (Figure 1), providing N=102 after standard-of-care venepuncture. Patients consented to clinical data access (Table 1).

**Table 1.**
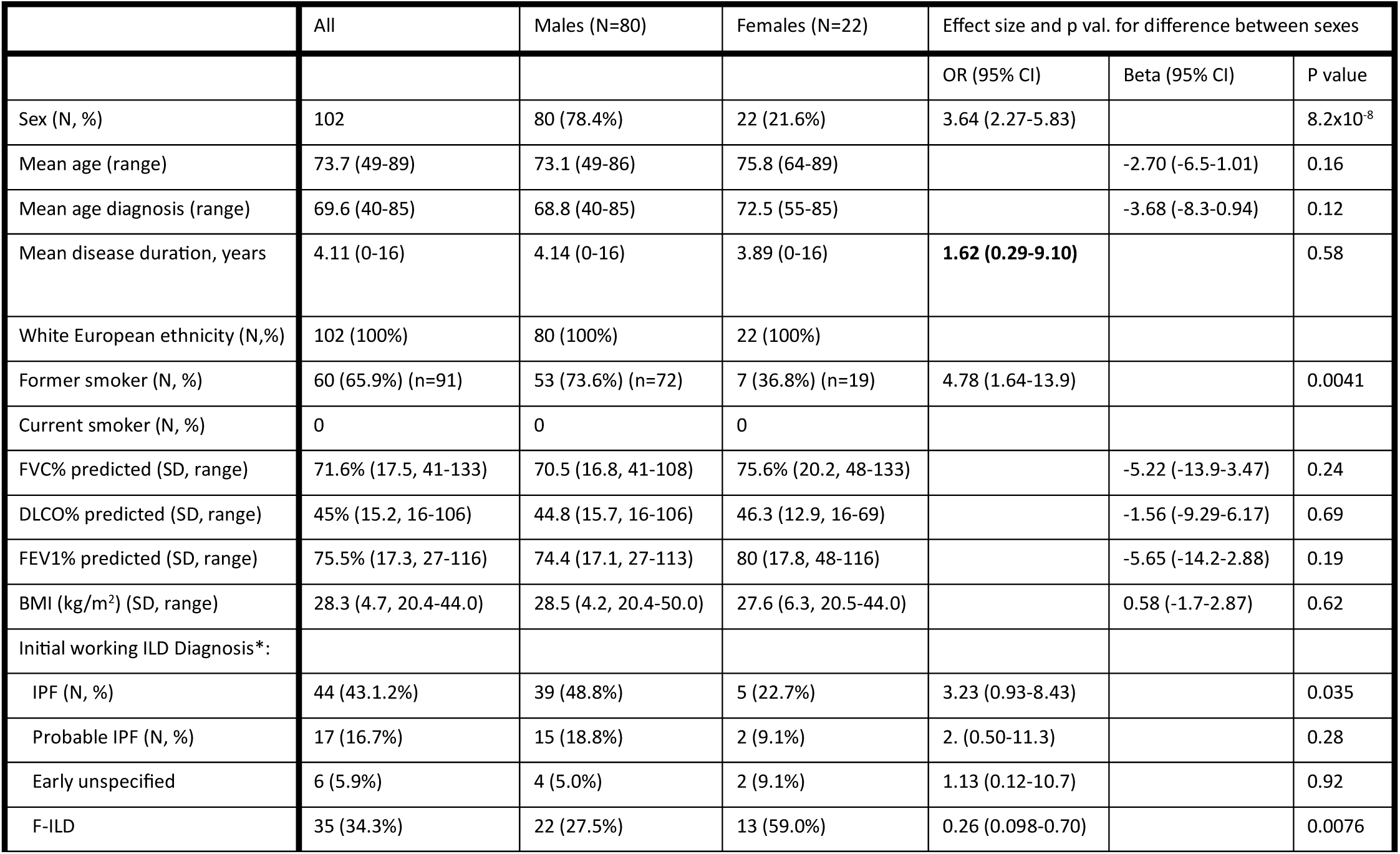
Demographics and baseline characteristics of the 102 patients enrolled in the study. Associations were adjusted for age (other than percent predicted means which already include adjustment). Lung function measures, FVC% (percent of predicted normal forced vital capacity), DLCO% (percent of predicted normal diffusing capacity for carbon monoxide) and FEV1% (percent of predicted normal volume exhaled in one second) were measured by clinical lung function tests, including spirometry. *Initial working diagnosis was corroborated at follow-up; all ‘probable IPF’ was confirmed as IPF and all ‘early unspecified’ was confirmed as non-IPF F-ILD, one patient initially diagnosed as IPF was re-diagnosed as RA-ILD at follow-up. Additional abbreviations, BMI: body mass index, ILD: interstitial lung disease, IPF: idiopathic pulmonary fibrosis, F-ILD: fibrotic-ILD.

|  | All | Males (N=80) | Females (N=22) | Effect size and p val. for difference between sexes |  |  |
| --- | --- | --- | --- | --- | --- | --- |
|  |  |  |  | OR (95% CI) | Beta (95% CI) | P value |
| Sex (N, %) | 102 | 80 (78.4%) | 22 (21.6%) | 3.64 (2.27-5.83) |  | 8.2x10<sup>-8</sup> |
| Mean age (range) | 73.7 (49-89) | 73.1 (49-86) | 75.8 (64-89) |  | -2.70 (-6.5-1.01) | 0.16 |
| Mean age diagnosis (range) | 69.6 (40-85) | 68.8 (40-85) | 72.5 (55-85) |  | -3.68 (-8.3-0.94) | 0.12 |
| Mean disease duration, years | 4.11 (0-16) | 4.14 (0-16) | 3.89 (0-16) | **1.62 (0.29-9.10)** |  | 0.58 |
| White European ethnicity (N,%) | 102 (100%) | 80 (100%) | 22 (100%) |  |  |  |
| Former smoker (N, %) | 60 (65.9%) (n=91) | 53 (73.6%) (n=72) | 7 (36.8%) (n=19) | 4.78 (1.64-13.9) |  | 0.0041 |
| Current smoker (N, %) | 0 | 0 | 0 |  |  |  |
| FVC% predicted (SD, range) | 71.6% (17.5, 41-133) | 70.5 (16.8, 41-108) | 75.6% (20.2, 48-133) |  | -5.22 (-13.9-3.47) | 0.24 |
| DLCO% predicted (SD, range) | 45% (15.2, 16-106) | 44.8 (15.7, 16-106) | 46.3 (12.9, 16-69) |  | -1.56 (-9.29-6.17) | 0.69 |
| FEV1% predicted (SD, range) | 75.5% (17.3, 27-116) | 74.4 (17.1, 27-113) | 80 (17.8, 48-116) |  | -5.65 (-14.2-2.88) | 0.19 |
| BMI (kg/m<sup>2</sup>) (SD, range) | 28.3 (4.7, 20.4-44.0) | 28.5 (4.2, 20.4-50.0) | 27.6 (6.3, 20.5-44.0) |  | 0.58 (-1.7-2.87) | 0.62 |
| Initial working ILD Diagnosis*: |  |  |  |  |  |  |
| IPF (N, %) | 44 (43.1.2%) | 39 (48.8%) | 5 (22.7%) | 3.23 (0.93-8.43) |  | 0.035 |
| Probable IPF (N, %) | 17 (16.7%) | 15 (18.8%) | 2 (9.1%) | 2. (0.50-11.3) |  | 0.28 |
| Early unspecified | 6 (5.9%) | 4 (5.0%) | 2 (9.1%) | 1.13 (0.12-10.7) |  | 0.92 |
| F-ILD | 35 (34.3%) | 22 (27.5%) | 13 (59.0%) | 0.26 (0.098-0.70) |  | 0.0076 |

There were no exclusions relating to standard of care treatment, sex, F-ILD subtype and all patients with successful venepuncture were included, to minimise restrictions for a future trial. Recruitment occurred March 2022 - February 2023. Participants provided written informed consent and could withdraw at any time. The study was approved by the Royal Devon & Exeter Tissue Bank (RDETB) steering committee (HRA-Research Ethics Service approval 21/YH/0159). Approval (19/SW/1059), for the use of age and sex-matched serum samples from the EXTEND (Exeter 10,000) Project biobank was received from the Peninsula Research Bank Steering Committee and NIHR Exeter Clinical Research Facility.

Sex specific questionnaires with input from clinicians and patients were used as a basis for participant interviews, with the purpose of increasing awareness of factors that may affect individual sex hormone levels (**Supplement S1**). All interviews were conducted by the same interviewer, by telephone. Follow-up analysis of participant survival from centralised death records was carried out in June 2025. Follow-up lung function data was unavailable.

Control samples were matched on sex, age (integer years) and ethnicity (**Table 2**). Controls with respiratory disease, chronic bronchitis/emphysema or a family history of either were excluded. Women taking hormone replacement therapy (HRT) and men taking testosterone were excluded. Information including medical history, smoking, BMI and self-reported age of menopause was available, plus stored serum and DNA samples.

**Table 2.** Demographics and baseline characteristics of age and sex-matched control group. Associations were adjusted for age. (BMI: Body Mass Index).

|  | All | Males | Females (N=22) | Effect size and p val. for difference between sexes |  |  |
| --- | --- | --- | --- | --- | --- | --- |
|  |  |  |  | OR (95% CI) | Beta (95% CI) | P value |
| Sex (N, %) | 102 | 80 (78.4%) | 22 (21.6%) |  |  |  |
| Mean age (range) | 73.7 (49-89) | 73.1 (49-86) | 75.8 (64-89) |  |  |  |
| White European ethnicity (N,%) | 102 (100%) | 80 (100%) | 22 (100%) |  |  |  |
| Former smoker (N, %) | 48 (47.1%) | 45 (56.3%) | 3 (13.1%) | 8.74 (2.37-32.2) |  | 0.0011 |
| Current Smoker (N, %) | 2 (2.0%) | 2 (2.5%) | 0 |  |  |  |
| BMI (kg/m<sup>2</sup>) (SD, range) | 27.5 (3.92, 18.8-40.3) | 27.4 (3.6, 19.8-36.7) | 27.7 (5.1,18.8-40.3) |  | -0.30 (-2.26-1.66) | 0.76 |

### Data Acquisition and Processing

Acquisition and processing of serum biomarker, leukocyte telomere length (LTL), lung function and questionnaire data are described in the **Supplement (S1)**. HT-STELA was chosen for its ability to measure absolute very short single telomere length with potential for high-throughput^19^.

### Statistical Analysis

Case-control analyses of means for the two groups were carried out using Mann-Whitney (MW) nonparametric testing using GraphPad Prism, Version 10.0.3 (217). Association analyses were carried out using linear and logistic regression and survival analysis was carried out using both Kaplan-Meier analysis and Cox regression, adjusting for covariates as described, using Stata/SE, Version 18.0.

### Comparative Analyses of UK Biobank Data

Additional sensitivity analyses to support the observations in a larger data set were carried out using the same statistical methods in the UK Biobank IPF and control datasets described previously^12^.

## RESULTS

### Safety characteristics

67/80 male and 17/22 female patients had Hb in the normal range, 63/80 male and 15/22 female patients had haematocrit in the normal range, 76/80 male and 17/22 female patients had both Hb and haematocrit below the upper limit of normal, which would facilitate potential testosterone treatment for the majority (**Figure 2**). Full blood safety results are detailed in **Supplement S2**.

**Figure 2:**
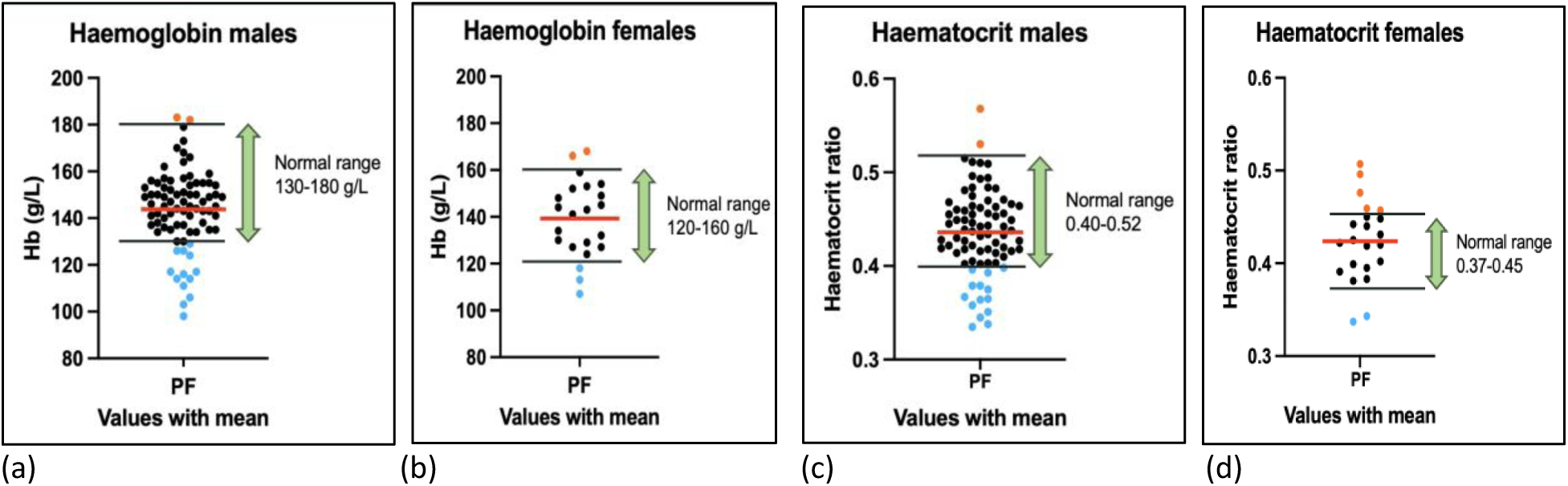
Haemoglobin and haematocrit values for male and female patients. (a) Hb for male STARSHIP patients, (b) Hb for female STARSHIP patients, (c) haematocrit for male STARSHIP patients, (d) haematocrit for female STARSHIP patients. Values determined by complete blood count.

### Telomere length

Mean leukocyte telomere length (LTL) measured using HT-STELA was shorter for N=101 ILD cases (4.57kb [95%CI: 4.46-4.69]) than for ASMCs (4.78kb [95%CI: 4.67-4.89]; p=0.0057; **Table 3** & **Figure 3a-c**). Additional telomere length results are described in **Supplement S3**.

**Table 3.** Leukocyte telomere length comparing cases and their individual matched controls.

|  | Mean LTL:<br>Cases (kb) | Range | 95%CI | N | Mean LTL:<br>Matched<br>Controls (kb) | Range | 95%CI | N | Mann-Whitney P |
| --- | --- | --- | --- | --- | --- | --- | --- | --- | --- |
| All | 4.57 | 3.41-6.52 | 4.46-4.68 | 101 | 4.78 | 3.69-6.43 | 4.67-4.89 | 98 | 0.0057 |
| Females | 4.49 | 3.95-6.15 | 4.24-4.74 | 22 | 4.73 | 3.95-6.15 | 4.50-4.96 | 22 | 0.196 |
| Males | 4.59 | 3.69-6.43 | 4.46-4.72 | 79 | 4.79 | 3.69-6.43 | 4.66-4.92 | 76 | 0.016 |
| IPF + probable IPF | 4.56 | 3.41-6.52 | 4.40-4.71 | 61 | 4.81 | 3.69-6.23 | 4.67-4.95 | 57 | 0.0063 |
| Early unspecified or<br>F-ILD | 4.59 | 3.63-6.02 | 4.42-4.76 | 40 | 4.73 | 3.74-6.43 | 4.55-4.91 | 41 | 0.28 |
| Familial PF | 4.47 | 3.41-5.72 | 4.15-4.80 | 17 | 4.85 | 4.22-6.15 | 4.62-5.09 | 17 | 0.093 |
| Sporadic PF | 4.59 | 3.49-6.52 | 4.45-4.73 | 74 | 4.79 | 3.69-6.43 | 4.65-4.92 | 71 | 0.035 |

**Figure 3:**
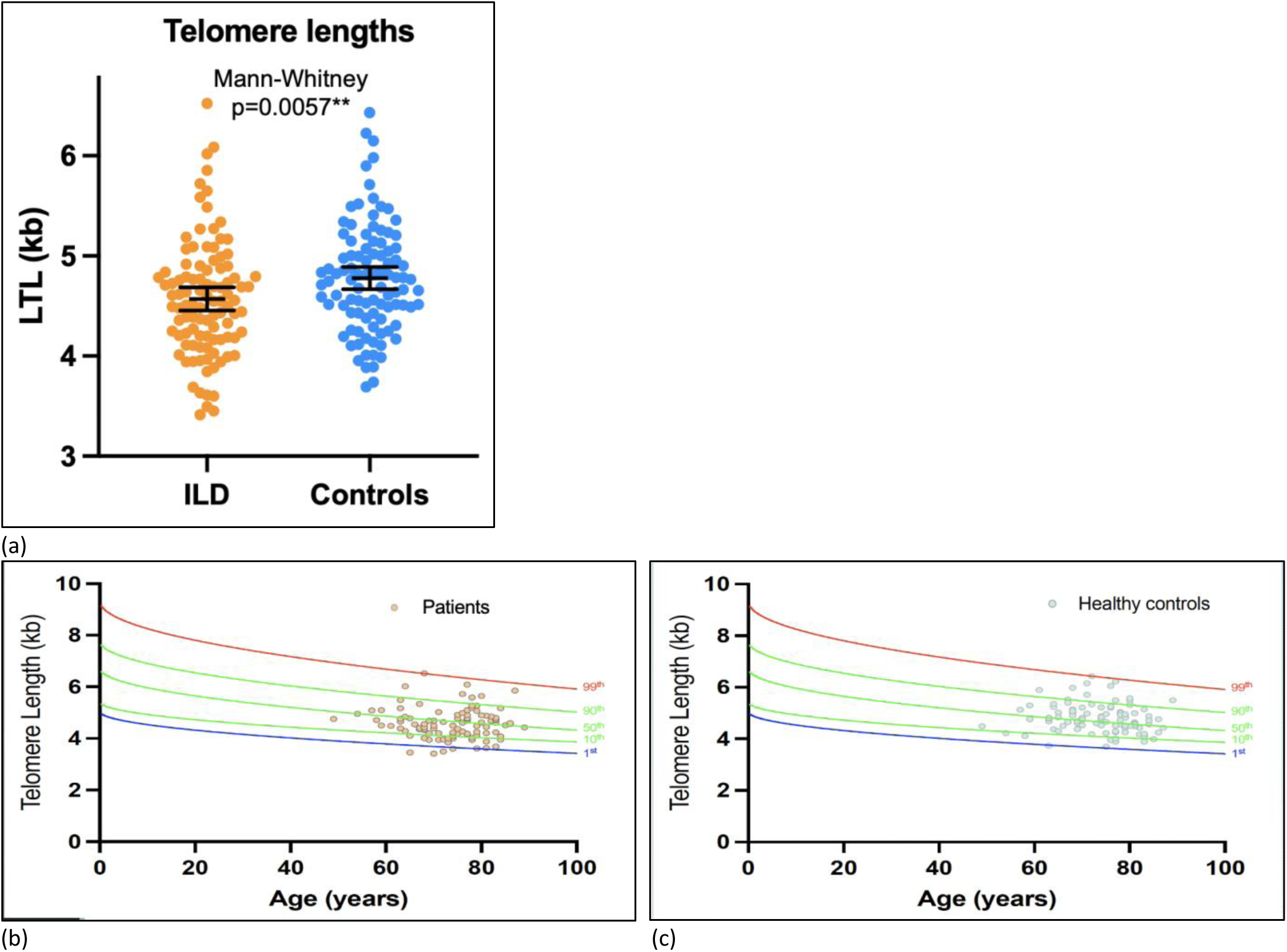
Telomere lengths measured using HT-STELA in PF patients and controls. (a) Leukocyte telomere lengths (LTL, showing mean, 95%CI and Mann-Whitney p-value) for 101 ILD patients and 99 age and sex matched controls, (b) LTL for PF patients plotted against existing STELA age centiles, (c) LTL for matched controls plotted against existing STELA age centiles.

### Endocrine results

Mean albumin concentration was lower for male patients than ASMCs and mean SHBG concentration was higher (**Table 4**, **Figure 4a**). Consequently, despite similar mean total testosterone, patients had significantly lower bioactive testosterone concentrations and percentages than ASMCs (where ‘bioactive’ represents bioavailable and free for males and free androgen index for females).

**Table 4.** Sex hormone concentrations and percentages for male PF patients and age-matched controls. Mann-Whitney P value is shown for comparisons between the two groups. One control sample with total testosterone below the measurement limit is assigned a value of 0.1nmol/L and highlighted with an asterisk*. (SHBG: sex hormone binding globulin, T: testosterone)

|  | Mean (SD):<br>Patients | Range | 95%CI | N | Mean (SD):<br>Controls | Range | 95%CI | N | Mann-Whitney P |
| --- | --- | --- | --- | --- | --- | --- | --- | --- | --- |
| Albumin (g/L) | 43.0 (3.0) | 36-49 | 42.4-43.7 | 80 | 44.2 (8.1) | 0-52 | 42.4-46.0 | 80 | <0.0001 |
| SHBG (nmol/L) | 53.2 (17.7) | 16.8-102 | 49.3-57.2 | 80 | 47.8 (20.4) | 19.1-140 | 43.2-52.3 | 80 | 0.0087 |
| Total T (nmol/L) | 14.0 (5.9) | 0.3-26.8 | 12.6-15.3 | 80 | 15.8 (7.0) | 0.1*-39.1 | 14.3-17.4 | 80 | 0.11 |
| Free T (pmol/L) | 211 (85) | 4-429 | 192-230 | 80 | 265 (100) | 0.6-514 | 243-287 | 79 | <0.0001 |
| Free T% | 1.56 (0.36) | 0.93-2.71 | 1.48-1.64 | 80 | 1.68 (0.41) | 0-2.68 | 1.59-1.78 | 80 | 0.0067 |
| Bioavailable T (nmol/L) | 4.95 (2.04) | 0.09-10.3 | 4.50-5.41 | 80 | 6.40 (2.59) | 0.01-13.7 | 5.82-6.98 | 79 | <0.0001 |
| Bioavailable T% | 36.6 (8.8) | 21.9-63.4 | 34.6-38.5 | 80 | 41.2 (10.4) | 5.6-63.8 | 38.9-43.6 | 79 | 0.0003 |

**Figure 4:**
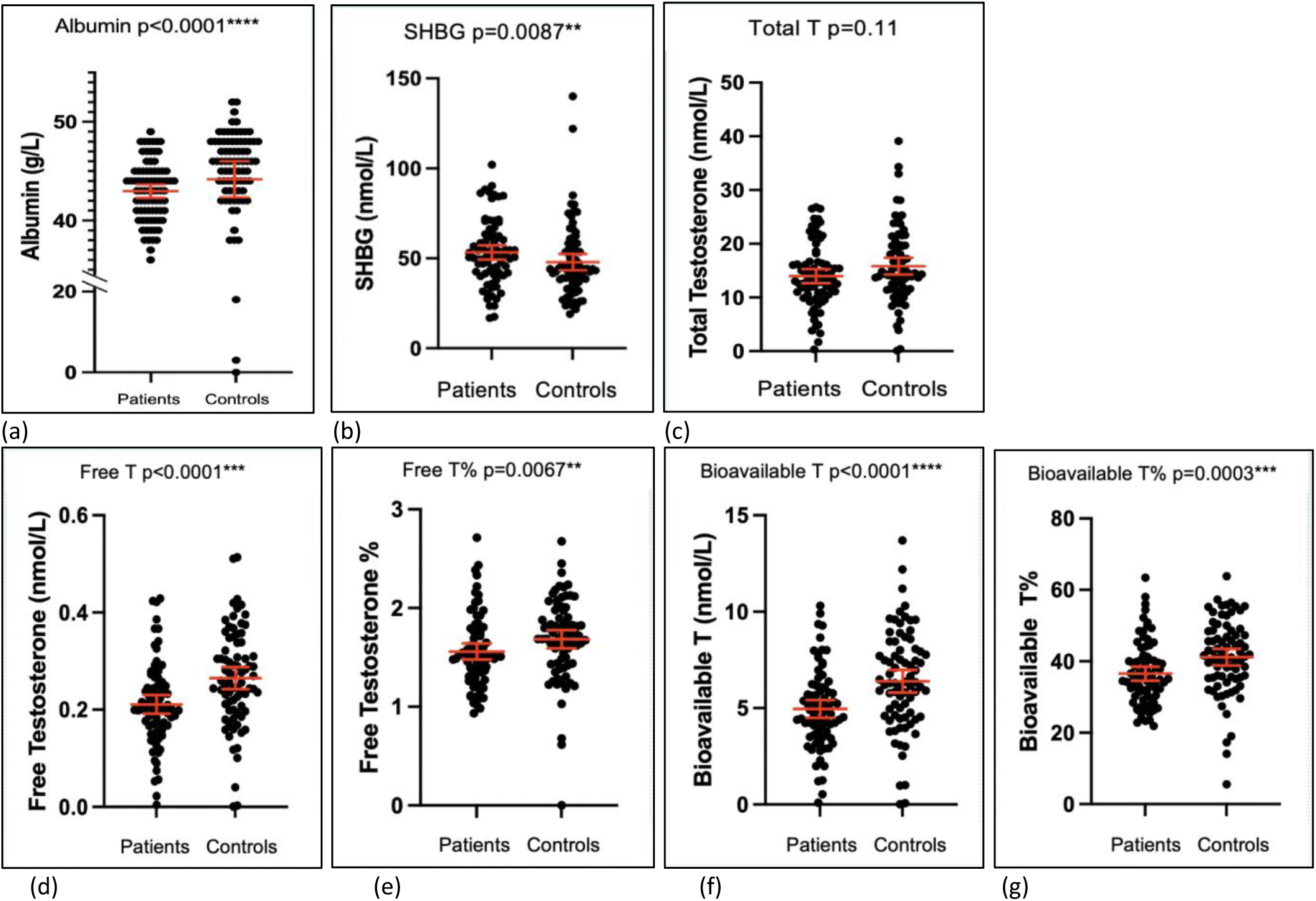
Scatter plots with mean and 95% confidence intervals and Mann-Whitney p values for sex hormones and related blood markers for N=80 male patients and age-matched controls. (a) Albumin concentrations,(b) Sex hormone binding globulin (SHBG) concentrations, (c) Total testosterone concentration, (d) Free testosterone concentration, (e) Free testosterone as a percentage of total testosterone, (f) Bioavailable testosterone concentration, (g) Bioavailable testosterone as a percentage of total testosterone.

Male patients had lower mean bioavailable testosterone concentration than ASMCs (4.95nmol/L [95%CI: 4.50-5.41] vs 6.40nmol/L [95%CI: 5.82-6.98]; p<0.0001, N=80 for both) and lower mean free testosterone (211pmol/L [95% CI: 192-230] vs 265 pmol/L [95% CI: 243-287]; p<0.0001). Indeed, the mean free testosterone for male patients was below the British Society of Sexual Medicine recommended testosterone replacement treatment threshold of 225pmol/L, with 50/80 (63%) at or below this value, compared with 25/80 (31%) ASMCs. No reduction of circulating testosterone by prednisolone treatment was observed (**Supplement S4**).

As for males, mean albumin concentration was lower for female patients than ASMCs and mean SHBG concentration was higher (**Table 5**, **Figure 5**). A UK Biobank study showed that low albumin precedes IPF disease onset rather than being caused by fibrosis (**Supplement S.5)**.

|  | Mean (SD):<br>Patients | Range | 95%CI | N | Mean (SD):<br>Controls | Range | 95%CI | N | Mann-Whitney P |
| --- | --- | --- | --- | --- | --- | --- | --- | --- | --- |
| Albumin (g/L) | 42.9 (2.7) | 38-48 | 41.7-44.1 | 22 | 44.7 (4.0) | 33-52 | 42.9-46.5 | 22 | 0.039 |
| SHBG (nmol/L) | 73.7 (34.1) | 24.8-179 | 58.6-88.8 | 22 | 52.9 (24.6) | 20.5-118 | 42.0-63.8 | 22 | 0.022 |
| Oestradiol (pmol/L) | 28 (15.4) | 18-61 | 21.2-34.8 | 22 | 30.5 (25.5) | 18-114 | 19.2-41.7 | 22 | 0.67 |
| Total T (pmol/L) | 395 (32) | 100*-1100 | 252-539 | 22 | 609 (529) | 100-1900 | 375-844 | 22 | 0.20 |
| Free Androgen Index | 0.51 (0.37) | 0.13-1.50 | 0.35-0.67 | 22 | 1.23 (1.04) | 0.14-3.93 | 0.77-1.69 | 22 | 0.0036 |
| Free Oestrogen index | 0.42 (0.25) | 0.16-1.34 | 0.31-0.53 | 22 | 0.75 (0.77) | 0.15-3.14 | 0.41-1.09 | 22 | 0.18 |

**Figure 5:**
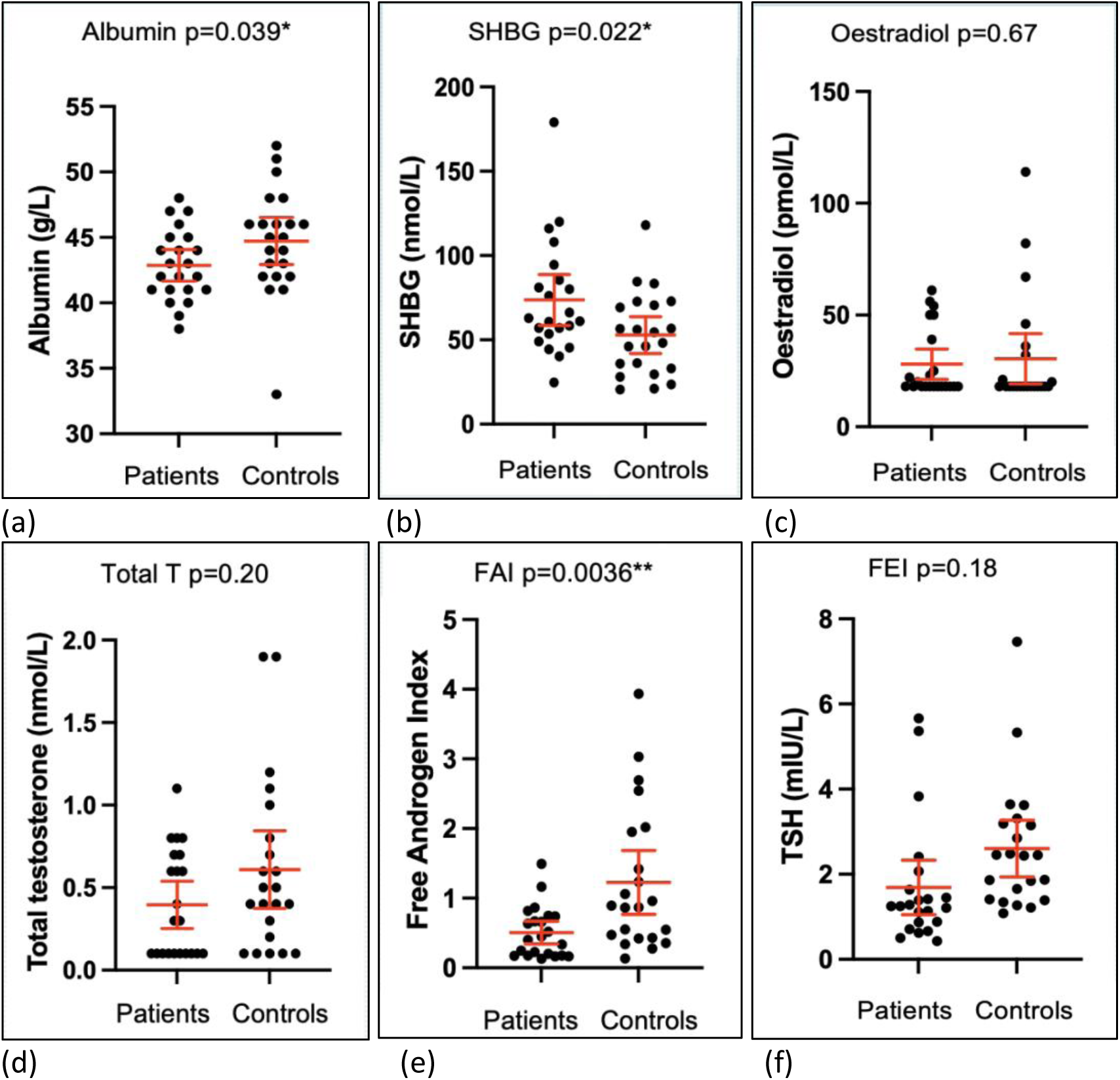
Scatter plots with mean and 95% confidence intervals and Mann-Whitney p values for sex hormones and related blood markers for N=22 female patients and age-matched controls, (a) Albumin concentrations (b) Sex hormone binding globulin (SHBG) concentrations, (c) Oestradiol concentration, (d) Total testosterone concentration, (e) Free Androgen Index, (f) Free Oestrogen Index.

Oestrogen concentrations were low for both female patients and controls, in keeping with post-menopausal age. In total, 12/22 patients and 14/22 controls had oestrogen values below the lower measurement limit of 19pmol/L. While only total oestrogen (i.e. bioavailable plus bound) is generally measured clinically, values of ‘free oestrogen index’^18^, FEI = (oestrogen concentration/SHBG concentration) were also calculated to account for the significantly higher SHBG values for patients. The mean was lower for patients than controls (0.42 [95%CI:0.31-0.53] vs 0.75 [95%CI:0.41-1.09]) but did not reach significance in available data.

Total testosterone in females is around 20 times lower than for males^20^. While only one male sample fell below the lower limit of total testosterone measurement of 0.2nmol/L, 10/22 (45%) of female patient samples and 5/22 (23%) of female control samples fell below the lower limit. We used an imputed testosterone median value between the detection limit and zero for these missing data, based on previous analyses in UK Biobank data^12^. The most appropriate clinical measure for testosterone in females, free androgen index, was lower for PF patients than ASMCs (mean FAI = 0.51 [95%CI: 0.35-0.67] vs 1.23 [95%CI: 0.77-1.69]; p= 0.0036).

Thus, in both male and female patients, data show lower albumin, higher SHBG and lower bioactive sex hormone levels in patients than ASMCs.

### Telomere length, sex hormone levels and lung function correlations

Strong associations between sex hormones and lung function were found. FVC% predicted was associated with free androgen index in female PF patients (**Figure 6**) and after adjusting for age and BMI, the linear regression association became stronger; *β*=34.3 (95%CI: 13.6-54.9), p=0.0029, N=20. Similarly, FVC% predicted was associated with bioavailable testosterone concentration in male patients; after adjusting for age and BMI, *β*=2.76 (95%CI: 0.97-4.56), p=0.0030, N=80. DLCO% predicted was also associated with bioavailable testosterone concentration in males; after adjusting for age, *β*=3.31 (95%CI: 1.60-5.02), p= 2.4×10^-4^, N=78. DLCO% predicted was not associated with free androgen index in the smaller female group.

**Figure 6:**
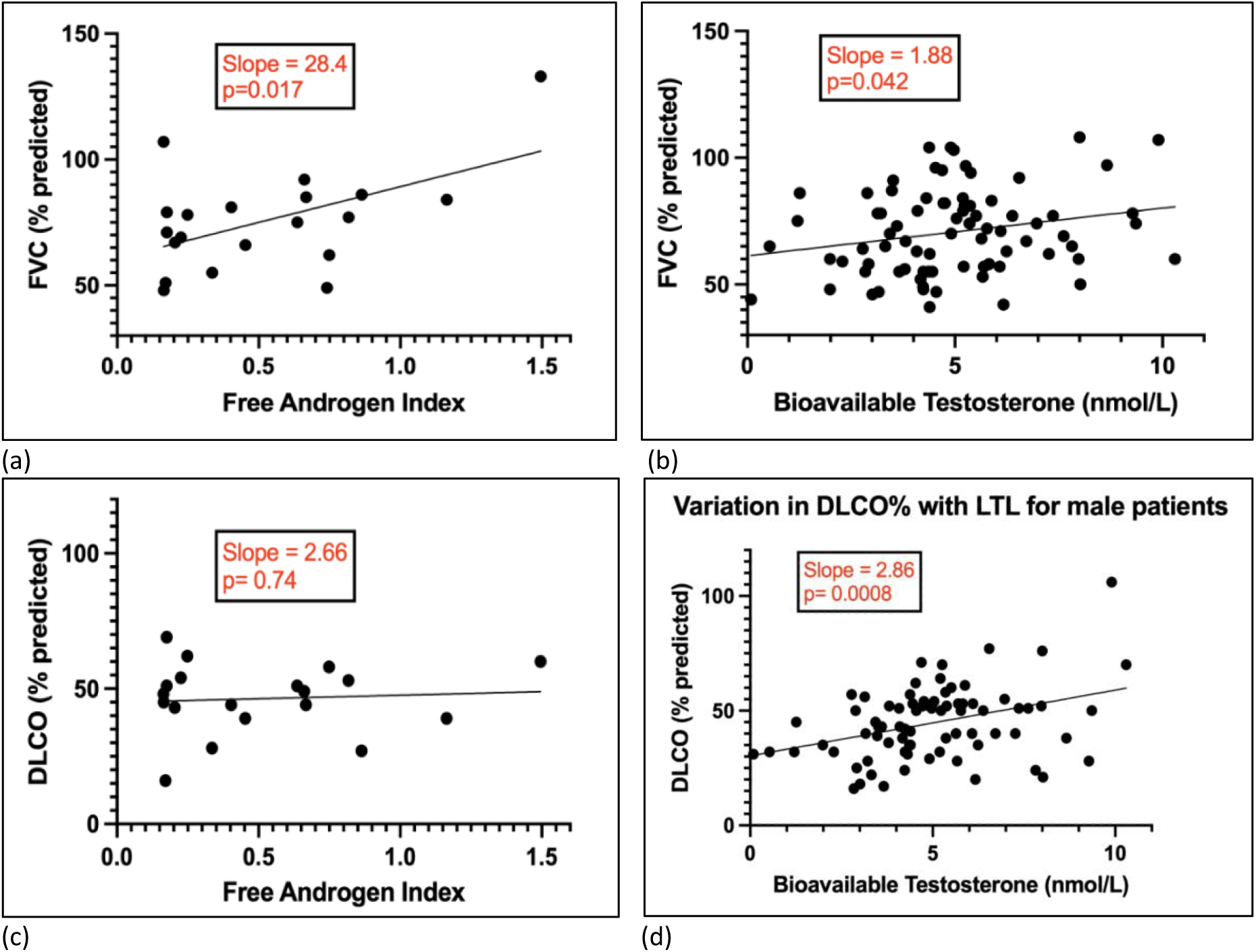
Association of lung function with the appropriate bioavailable testosterone measures in female and male PF patients. (a) Association of FVC% predicted with free androgen index in n=20 female PF patients, (b) Association of FVC% predicted with bioavailable testosterone concentration in n=80 male PF patients, (c) No association between DLCO% predicted and free androgen index in n=20 female PF patients, (d) Association of DLCO% predicted with bioavailable testosterone concentration in n=80 male PF patients. Plotted associations were derived using linear regression and contain no age adjustment.

Weak associations were found between telomere length and sex hormones and no associations were seen between lung function and telomere length (**Supplement S6**), where the impact of smoking on sex hormones and LTL is also described.

### Survival

During the period from study recruitment (March 2022 - February 2023) to census (June 2025), 51/102 (50%) of the STARSHIP study patients died. Median follow-up time was 33 (28-39) months. Of the male patients, 31/50 (62%) with low and 9/30 (30%) with normal free testosterone had died; Kaplan-Meier analysis demonstrated a significant survival difference between the two groups (log rank test Chi^2^=6.8, p=0.0092, N=80) and even more extreme difference when the group was split into three, **Figures 7a,b**,. In Cox proportional hazard modelling adjusted for key confounders, (baseline age and lung function at presentation; both FVC and DLCO), low free testosterone (<225pmol/L) was associated with increased risk of death (HR=2.23, p=0.038, N=78; **Supplement S7**). This was strengthened by inclusion of additional confounders (IPF diagnosis, LTL, antifibrotic and immunomodulatory treatments); HR=2.66, p=0.023, N=77). Neither IPF diagnosis nor LTL alone reached significance in these models. When comparing definite IPF vs non-IPF F-ILD cases, the proportion of patients with low free testosterone was slightly higher in the non-IPF group (23/31=74% vs 27/49=55%, Chi^2^ p=0.086).

**Figure 7:**
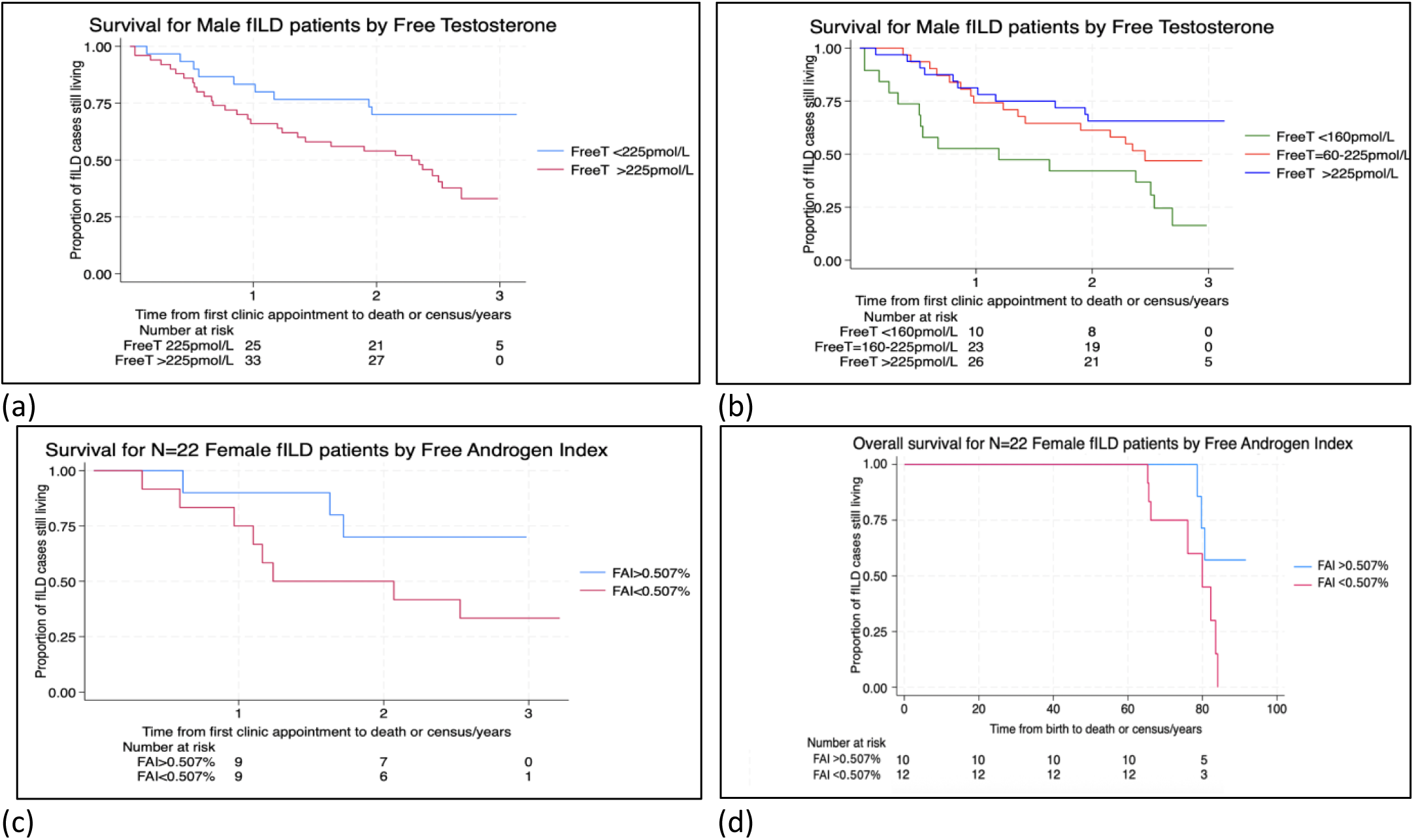
Kaplan-Meier survival curves for male and female pulmonary fibrosis patients split by bioactive testosterone, (a) Survival for male patients by free testosterone concentration above and below 225pmol/L at study consent (b) Survival for male patients by free testosterone concentration below 160pmol/L, between 160-225pmol/L and above 225pmol/L at study consent (c) Survival for female patients by free androgen index above and below mean FAI =0.507% at study consent, (c) Overall survival for female patients by free androgen index above and below mean FAI =0.507% (p=0.046) at study consent (no corresponding difference was seen for males).

Of the female patients, 8/12 (67%) with below and 3/10 (30%) with above mean free androgen index for the group (FAI=0.507%) had died by the census date; Kaplan-Meier analysis suggested a survival difference between groups although this did not reach significance with N=22 (Chi^2^=2.7, p=0.10), **Figure 7c**. However lower FAI was associated with worse lifetime survival (Chi^2^=4.8, p=0.029, **Figure 7d**) with a mean survival advantage of 6.56 years (95%CI:0.13-13.0, p=0.046) for the higher FAI group amongst deceased females. This is particularly notable since 6/10 patients in the higher FAI group also have clinically low FAI (<0.8), suggesting additional treatment benefit. No corresponding survival advantage was observed for free testosterone in males (p=0.47). In Cox proportional hazard modelling for females adjusted for age, FVC and DLCO, low free androgen index (<0.507%) did not show increased mortality risk from diagnosis with this small sample size (HR=3.59, p=0.22, N=18), nor were significant associations apparent in the more comprehensive model (**Supplement S7**). When comparing definite IPF vs non-IPF F-ILD females (N=18), the proportion with below mean FAI was again biased towards the non-IPF group (9/14=64% vs 3/8=38%, Chi^2^ p=0.23) although this did not reach statistical significance.

### Interview findings

Reported features suggesting possible hormonal imbalances were common amongst this patient group and are highlighted for 47/74 (63.5%) males and 17/19 (89%) females for whom interviews were conducted (Figure 8).

**Figure 8:**
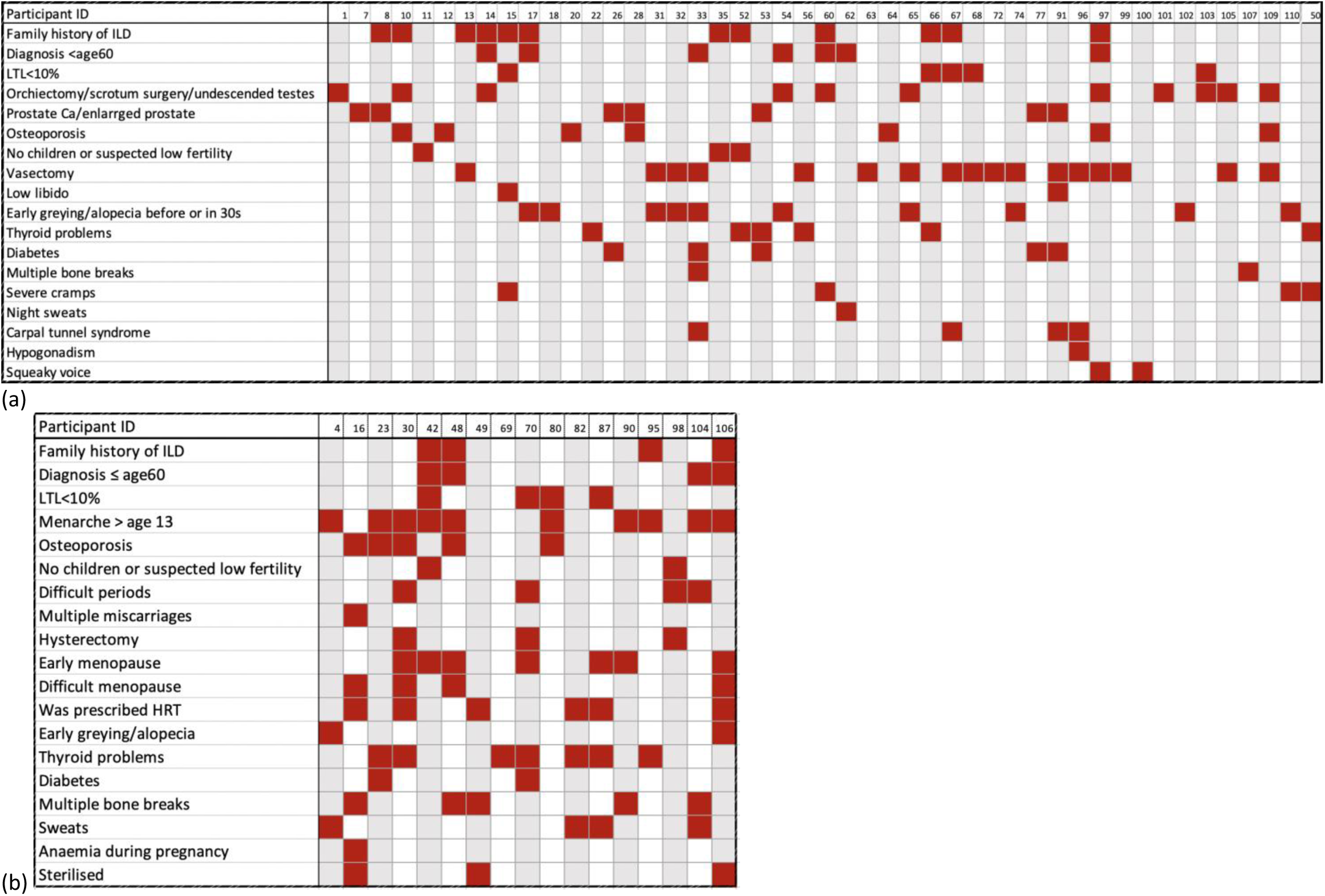
Symptoms reported in interview that may relate to low sex hormone levels in (a) Males and (b) Females with PF.

As additional observations that may or may not be relevant here, one reported a history of undescended testis and two reported undescended testes. Two reported single orchidectomy. Hair loss/early balding have been included here, along with early hair greying, because both may have both a genetic and hormonal component and comments suggested that males from families with PF often presented with either early greying or early balding. Excerpts from feedback from two female patients highlighting the kinds of symptoms reported are included in Supplement S.8.

Standard questionnaire data linked with the control samples allowed a direct comparison for some parameters. For females, mean reported age of menarche was 13.5 (range 11-17) for the 19 patients interviewed, compared to mean age 13.0 (range 11-16) for 20 controls (p=0.33). Mean age of first menopause symptoms for 14/19 patients who knew was 45.6 (range 35-52) and for 16/22 controls who knew, mean age of first menopause symptoms = 50.2 (range 42-60), p=0.018.

In addition, 16 (17.2%) respondent patients (8M=11% : 8F=42%) self-reported being prescribed levothyroxine, compared with no ASMCs (Chi2 p<0.0001), 14 (15%) (9M=12%, 5F=26%) patients had previously been prescribed Alendronic acid for osteoporosis compared with 5 matched controls (Chi^2^ p=0.027) and a further 5 patients (2M) reported a history of multiple bone fractures without reference to osteoporosis. 7/19 (37%) female patients had received HRT compared with no female controls. This finding stands out because the control group was recruited earlier than the patient group (during the period 2010-2023), and HRT was more readily prescribed pre-2002 when more of the control group would have been pre-menopausal.

In summary, qualitative findings suggest a medical history in which sex hormone insufficiency may play a role.

## DISCUSSION

Previously we reported findings suggesting that sex hormone replenishment may protect against PF onset and progression, possibly by slowing critical telomere shortening for those at risk^12^. While hypoxaemia can reduce testosterone^21^ and the question of causality is an important one, there we also reported both genetic and *in vitro* mechanistic studies suggesting that sex hormones influence telomere length. In fact, we suspect bidirectional causality between telomere length and sex hormones/SHBG and draw on similar findings in type II diabetes (see below) where treatment of testosterone insufficiency improves glycaemic index and survival^22,23^. The question of treatment benefit requires a randomised PF/control trial, with baseline safety information from a representative patient cohort. This study demonstrated that baseline haemoglobin and haematocrit levels were within normal range for most male patients, making testosterone treatment possible.

Perhaps surprising for a pragmatically-sized study, we have shown consistent statistically significant evidence of lower levels of sex hormones amongst patients, compared with ASMCs. Of particular note was the high proportion of male patients (63%) with low free testosterone (<225pmol/L), compared with 31% of ASMCs. Concentrations of albumin were lower and SHBG higher than for ASMCs, leading to lower bioavailability of sex hormones (demonstrated for testosterone in both sexes). Lung function (FVC% and DLCO%) was associated with bioavailable testosterone concentration in males and FVC% was associated with FAI in females. Longitudinal follow-up showed substantially poorer survival for patients of both sexes with lower bioactive testosterone.

We have shown previously that IPF prevalence amongst males associates with low bioavailable testosterone/high total testosterone, due to high circulating SHBG^12^. This study shows higher SHBG for both male and female PF patients compared with age-matched controls, suggesting that the Vermeulen equations used for oestradiol^24^ or the ‘free oestrogen index’ described earlier^18^ might be helpful in future for monitoring oestrogen bioavailability clinically. In this age-group for post-menopausal women, total oestrogen levels are low and difficult to measure. High SHBG concentrations might act as a useful marker for low free oestrogen^18^. Strong evidence supporting testosterone treatment for both male and female PF patients who have low bioavailable testosterone is provided in Supplement S.9.

No association was observed between lung function and telomere length. Mean telomere length in the blood declines slowly with age but changes very little throughout life for an individual; the heritable fraction of leukocyte telomere length is 70%^25^. The reported association of leukocyte telomere length with survival in PF^26^ is more likely an association with strength of genetic predisposition to shorter telomeres than a measure of disease progression and still has importance for clinical management (See Supplement S.10 regarding inferences for telomere length testing in F-ILD).

This STARSHIP study has several limitations. Typical for the South West UK, all patients (and hence controls) were of white ethnicity. Only 22/102 (21.6%) were female which and, typical for this patient population, all were post-menopausal. Further studies involving female patients are needed to boost statistical power to produce significant results for women. For 12/22 (55%) female patients and 14/22 (64%) age-matched controls, oestradiol concentrations were below the NHS laboratory measurement limit. This is typical for post-menopausal women (of whom there are approximately 11 million in the UK^27^). Similarly, testosterone is best measured by mass spectrometry and pre-sample fasting was not undertaken by patients or controls. Measurements for 10/22 (45%) female patients and 5/22 (23%) female controls were below the measurement limit. The comparison of interview data was limited by what had been recorded as standard for the ASMC dataset. More probing questions could potentially have been asked about additional issues relating to sex hormone levels such as libido and infertility. Sexual dysfunction has since been reported in PF^28^.

The study findings present a picture of lower bioavailable oestrogen and testosterone amongst patients with pulmonary fibrosis that importantly seem to associate with lung function and survival. The prospect of treating PF patients with sex hormones appears to be relatively safe and has an established precedent in diabetes (**Supplement S8**). Indeed, recent evidence suggests that testosterone improves lung health generally in both sexes and that high SHBG may be detrimental^29^. Patient members of our research group Exeter Patients in Collaboration for Pulmonary Fibrosis Research or ‘EPIC PF’ voiced their unreserved support for sex hormone therapy treatment trials. These data provide support for well-designed randomised-controlled trials for patients that may alleviate symptoms, improve wellbeing and quality of life. They may also play a future role in delaying or preventing onset of disease for family members with short telomeres.

## Supporting information

STARSHIP Supplement

## Data Availability

All data produced in the present work are contained in the manuscript

## ACKNOWLEDGEMENTS

Initial results from this study were presented at the American Thoracic Society Conference 2024^30^ and received an invited talk at the European Respiratory Society Congress 2025. The longitudinal results were presented at the British Thoracic Society Winter Meeting in 2025^31^ and highlighted as an ‘emerging insight’ in the Conference Review^32^.

This study was supported in part by grant MR/N0137941/1 for the GW4 BIOMED MRC DTP, awarded to the Universities of Bath, Bristol, Cardiff and Exeter from the Medical Research Council (MRC)/UKRI. Additional funding was provided by Royal Devon & Exeter NHS Trust Research Capacity Funding. JKP is supported by the UKRI Expanding Excellence in England award. CJS is supported by MRC project grants (MR/V002538/1 and MR/S002626/1). This UK Biobank component of this research has been conducted using UK Biobank Resource (applications 9072 and 44046).

The National Institute for Health and Care Research (NIHR) Exeter Clinical Research Facility is a partnership between the University of Exeter Medical School College of Medicine and Health, and Royal Devon University Healthcare NHS Foundation Trust. This project is supported by the NIHR Exeter Clinical Research Facility. The views expressed are those of the author(s) and not necessarily those of the NIHR or the Department of Health and Social Care.

## COMPETING INTERESTS

DMB and KN are shareholders of the Cardiff University spin out company Telonostix Ltd that generated the telomere length data for this study.

## Notes

### Author Declarations

Royal Devon & Exeter Tissue Bank (RDETB) steering committee gave ethical approval for this study (HRA - Research Ethics Service approval 21/YH/0159). Peninsula Research Bank Steering Committee and NIHR Exeter Clinical Research Facility gave approval for access to EXTEND Project Biobank for control samples, Approval (19/SW/1059),

