## Supplementary material for "STARSHIP: Study of Telomeres And Role of Sex Hormones In Pulmonary fibrosis": STARSHIP Supplement

**Supplementary Information**

#### S.1 Blood biomarker, telomere length, lung function and questionnaire data acquisition and processing

##### S1.1 Overview

Leukocyte telomere length was measured using high-throughput single telomere length analysis, HT-STELA[1] (Supplement S.2). This method amplifies the double stranded region of telomeres using polymerase chain reaction (PCR) at specific chromosome ends (usually the 17p telomere). HT-STELA was chosen as the measurement method because it measures absolute kilobase pairs (kb) and can measure the very short telomere lengths thought to be important in PF. All 204 anonymised samples were analysed as a single batch and results plotted against normal telomere length age centiles.

Similarly, 204 stored serum samples underwent batch analysis of: testosterone concentration, T (nmol/L); sex hormone binding globulin, SHBG (nmol/L); albumin (g/L); oestradiol (pmol/L; females only); and thyroid stimulating hormone, TSH (mIU/L). Assay technical information is included in Supplement S2. From these values, free and bioavailable testosterone values were calculated for males (using the Vermeulen equation[2]) and free androgen index (FAI) values for females (using the equation: FAI=(T/SHBG)*100).

Clinical data, including lung function measures (forced vital capacity [FVC] and diffusing capacity in the lung for carbon monoxide [DLCO]) and clinic day blood test results (Full blood count (FBC, including haemoglobin (g/L; normal range male: 130-180g/L, normal range female: 120-160g/L), haematocrit (ratio; normal range male: 0.40-0.52, normal range female: 0.37-0.45)) as part of routine, standard care, medications and MDT diagnosis were extracted from patient records with consent and were anonymised.

As a secondary aim of the study, two gender-specific questionnaires with input from clinicians and patients were used as a basis for participant interviews (Supplement S.2). All interviews were conducted by the same interviewer (AD), by telephone. Verbatim questionnaire responses were gathered by the lead researcher (AD) from telephone interviews with 93 patients. The data were separated into quantitative numerical data and qualitative subjective verbal responses and reviewed independently by a second researcher (AMR).

##### S.1.2 Blood sample processing details

**Two blood samples** were taken for STARSHIP, together totalling no more than 10ml, one in an EDTA tube (A) and the other in a serum gel tube (B). The samples were then processed and stored as follows:

**A) The EDTA sample (lavender top tube, 4ml)**

(i) Gently invert tube 5-10x after collection to mix in the anticoagulant and centrifuge within 2hrs.
(ii) Centrifuge sample at 1400*g* and 4°C for 10 minutes
(iii) Transfer the plasma tubes into labelled microcentrifuge tubes, taking care not to disturb the buffy coat
(iv) Aspirate the buffy coat layer into a labelled tube for freezing and batch transfer to Cardiff for STELA (address below)*
(v) Retain the red blood cell aliquot in a labelled tube for freezing
(vi) Centrifuge the plasma in the microcentrifuge tubes at 13,000rpm for 10 minutes.
(vii) Aliquot 1ml of plasma into a labelled cryo tube and repeat as necessary. Take care not to disturb the pellet at the bottom of the microcentrifuge tube.
(viii) Store cell free plasma, buffy coat sample and red cell sample immediately in the appropriate rack and location in -80°C freezer.

B) **The serum separator sample (gold top tube, 4ml)**

(i) Gently invert tube 5-10x after collection to mix in the gel
(ii) Allow sample to clot for 30 minutes at room temperature and centrifuge within 2 hours.
(iii) Centrifuge sample at 1400*g* and 4°C for 10 minutes
(iv) Aspirate the serum into a labelled tube suitable for later batch analysis in the RILD clinical laboratory
(ix) Store serum (for sex hormone panel) and labelled tube of cellular components (for future studies) immediately in the appropriate rack and location in -80°C freezer.

##### S.1.3 HT-STELA and Sex Hormone Analysis

Leukocyte telomere length for both cases and controls was measured using high-throughput single telomere length analysis, HT-STELA[1] , at TeloNostiX Ltd (Cardiff, UK). This method utilises the polymerase chain reaction (PCR) to amplify the double stranded region of telomeres at specific chromosome ends (usually the 17p telomere); amplified products are detected with capillary electrophoresis using a Fragment Analyzer system (Agilent) and quantified using the ProSize data analysis software (Agilent).

HT-STELA was chosen as the optimum measurement method for STARSHIP participants because it:

**(i)** measures absolute in kilobase pairs (kb) rather than relative telomere length compared with a reference (making it more useful for comparisons),

**(ii)** can measure the very short telomere lengths thought to be important in determining cell fate in PF,

**(iii)** is not constrained by a lower limit of telomere length detection common to hybridisation-based methods such as terminal restriction fragment analysis (TRF) and FISH,

**(iv)** Is DNA-based so does not require viable cells (flowFISH) or cells that are capable proliferation to provide metaphase spreads for quantitative in-situ hybridisation (Q-FISH),

**(v)** is robust to measurement variation (HT-STELA displayed a low measurement error with inter- and intra-assay coefficient of variance of 2.3% and 1.8%, compared with reported CVs of 6-20% for Q-PCR based methodologies) which makes it useful for sampling for individuals (rather than large-analysis such as UK Biobank)

**(vi)** is suitable for longitudinal measurements due to the low measurement error

**(vii)** has been designed for high throughput and has ISO17025 lab quality assurance accreditation,

(viii) is in routine use for the diagnosis of telomere biology disorders.

Following venepuncture, 102 STARSHIP patient peripheral blood samples were spun from EDTA tubes on the day of draw, aliquoted to extract the buffy coat layer which was frozen for batch analysis and shipped as frozen buffy coat samples. DNA was extracted from buffy coats samples using the Promega Maxwell system with the Maxwell® RSC Blood DNA Kit (Promega, USA). 102 matched control DNA samples were obtained from the EXTEND biobank; HT-STELA requires only around 50ng of DNA per analysis[1].

Similarly, 102 STARSHIP patient serum samples were frozen on the day of draw and stored for batch analysis, alongside 102 control serum samples from the EXTEND biobank. All 204 samples were transferred to the Exeter Clinical Laboratory International for batch analysis of: testosterone concentration, T (nmol/L); sex hormone binding globulin, SHBG (nmol/L); albumin (g/L); oestradiol (pmol/L; females only); and thyroid stimulating hormone, TSH (mIU/L). Samples were analysed for the measurement of T, and oestradiol using automated chemiluminescent immunoassays (Abbott Diagnostics). Reference ranges were as follows: T, 10–28 nmol/L; oestradiol less than 190 pmol/L. The respective intra-assay and inter-assay coefficients of variation for each assay were 4.2% and 2.8% (total T); and 3.3% and 3.0% (oestradiol). Analytical sensitivities were: 2 nmol/L (total T), and 37 pmol/L (oestradiol) From these values, free and bioavailable testosterone values were calculated for males (using the Vermeulen equation[2]) and free androgen index (FAI) values for females (using the equation: FAI=(T/SHBG)*100 ).

**S1.4 STARSHIP questionnaires for Females and Males**

The questionnaire proformas for interviews with female and male patients respectively are contained in the next 14 pages. The questionnaire was designed to gather general background information (14 questions), information about birth (5 questions), early symptoms of pulmonary fibrosis (3 questions), general medical history (8 questions for males and 7 for females), family history (8 questions), hormone history (5 questions for males and 14 for females) and additional questions (5). The questions were composed by the lead researcher (AD) with detailed expert input from clinicians (JP, AMR) and volunteer patient members of the Exeter Patients in Collaboration for PF research or 'EPIC PF' group. Some questions were constructed to enable direct comparison with data available for UK Biobank participants. Patients were given a copy of the patient questionnaire in their ILD clinic patient information pack and consented to be contacted by telephone for a follow up interview based on the questionnaire. Since most patients received their ILD diagnosis at this clinic and had a lot of difficult information to process, a period of several weeks was allowed before the researcher contacted them for interview. Three patients died prior to interviews being arranged. A further 6 patients could not be reached by telephone or opted not to undertake interview when contacted. A total of 93 patients re-confirmed their consent to interview.

Questionnaires were printed on differently coloured paper for M and F to ensure each patient was given the correct version and this made them easy for patients to find amongst the pack of information given to them at the outpatient ILD clinic. Patients consented on the day of their clinic visit to being contacted by telephone for a follow up interview based on the questionnaire. They were not asked to complete the questionnaire and were told that the questionnaire was just for information (a small number of patients chose to complete it anyway and sent it in to the ILD office prior to being contacted).

Patients were contacted by telephone and phone or text messages by the same interviewer/ researcher (AD) throughout, and contact details with a personal message were left for them if they were unavailable. Calls were made either late morning or early to mid-afternoon so that patients would not be disturbed at times when they might be more likely to be resting. Many said they were happy to conduct the interview at the time of the call and alternative times were arranged if the time of the call did not suit them.

The discussion was started by the interviewer asking how they were doing and listening carefully and empathetically to what they said and how they sounded. If they were well enough, they were then asked whether they still wished to be interviewed over the phone for the STARSHIP study for which they had consented while attending the ILD clinic a few weeks earlier. Several patients described problems or upsets that they had experienced since their appointment and the researcher was able to support them in getting issues resolved via clinical team.

Interviews were deliberately not recorded in order to retain the private and personal nature of the conversation, similar to a clinical consultation. The interviewer was a qualified coach who had conducted >1000 confidential coaching sessions with clients and was experienced in capturing accurate and detailed information from conversations. Verbatim patient comments were recorded on the questionnaire forms as the discussion unfolded. The questionnaire served as a guide but the patient was able to talk freely and share any concerns.

All of the female patients who agreed to be interviewed answered the questions themselves. One male patient suffered from Alzheimer's, was living in a care home and his son volunteered to answer the questions on his behalf. Another had poor hearing and his wife answered most of the questions, consulting him as necessary. A third was too breathless and tired so his wife answered the questions. Several of the male patients consulted their wives during the interview or answered the questions together. Consistency across interviews was maintained due to having a single interviewer for all 93 interviews.

Raw interview data was collected onto each of the 93 electronic questionnaire documents and stored securely. It contains the conversational content and is available for reference. The content deemed to have relevance to the topics of interest (sex hormone interactions reported here and evidence suggesting familial pulmonary fibrosis reported later) was transferred to a spreadsheet for analysis. Data was analysed by the interviewer/researcher and observations possibly relating to sex hormones were captured.

The average interview time per patient was approximately 30 minutes with range approximately 15 minutes (for a particularly breathless patient, who was nonetheless keen to contribute) to over an hour for a small number of patients who wanted to share in depth. The response to participation was very positive, with a sense that the timing and manner of the discussion created an opportunity for helpful discussion. Many patients expressed gratitude for the conversation.

### Study of Telomeres and Role of Sex Hormones in PF ('STARSHIP') - **Female Patient** Extended Clinical Review Form

The STARSHIP study has been set up to explore the idea that sex hormone therapy might be useful as a treatment option for patients diagnosed with pulmonary fibrosis. Completion of this form will help us to understand this.

**Please answer these questions as fully as you can and are happy to do so.**

| **General questions** | |
|---|---|
| Your unique study identifier | |
| Designated sex at birth | Male___ Female___ |
| Your age (years) | |
| Today's date (dd/mm/yyyy) | |
| Your postcode *(we use this to calculate things like local pollution level)* | |
| Have you ever smoked? *(please tick or highlight your choice)* | No___ Yes, previously__ Yes, currently__ |
| If yes, in what year did you start smoking? | |
| If yes and you have stopped, in which year? | |
| What did/do you smoke? *(please tick/highlight all that apply)* | Pre-rolled cigarettes____<br>Roll my own cigarettes ___<br>Cigars___<br>Pipe___<br>Vape___<br>Other (please specify)________________ |
| On average, how much did / do you smoke per day (please include grams of tobacco if you know that)? | |
| In which of these bands does your average total household income fall? *(please circle/highlight), (we use this to compare your data with that from our initial study on participants in the UK Biobank).* | 1) <£18000 (2) £18000-£30999<br>(3) £31000-£51999 (4) £52000-£100000<br>(5) >£100000. |
| Do you exercise? *(please tick/highlight)* | Yes___ No___ |
| What type of exercise do you do on a weekly basis? *(please tick/highlight all that apply)* | Yoga___ gentle walking___ brisk walking___<br>cycling___ Pilates___ running ___<br>gardening__ golf__ bowling__ swimming__<br>pulmonary rehabilitation exercises__<br>housework___ other: |
| How long do you spend exercising in an average week? | |
| Do you drink alcohol? | Yes___ No___ |

On average, what alcohol and how many units of each do you drink per week?

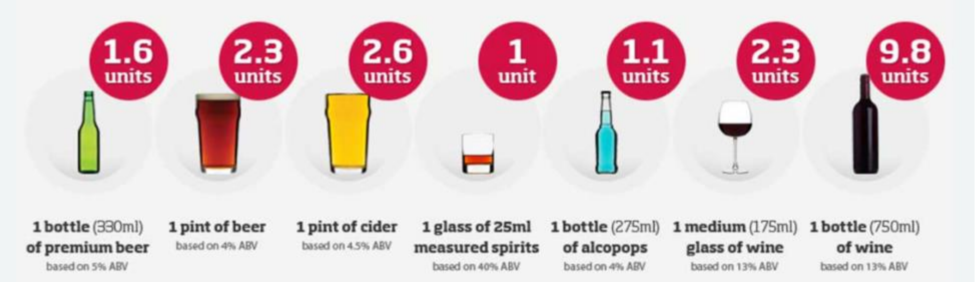

| | |
|---|---|
| Are you retired? | Yes___ No___ |
| What is/was your main occupation(s)? | |
| If you are retired, at what age did you retire? | |
| **Questions about your birth** | |
| To the best of your knowledge, were you born at full term, e.g. between 38-42 weeks of your mother's pregnancy? | Yes___ No___ |
| If no, were you born prematurely (i.e. before 'term' at 37 weeks)? | Yes___ No___ |
| If you were born before full term at how many weeks of your mother's pregnancy were you born? | ________ weeks Not sure |
| What was your approximate birthweight? | ___ lb___oz Don't know____ |
| Do you have any additional comments that you would like to add to any of your answers above? | |

| **Pulmonary Fibrosis** |
|---|
| How old were you when your PF was diagnosed? |
| Looking back, at what age do you think you had first signs of PF? |

| | |
|---|---|
| What were the early signs and symptoms? | Breathlessness___<br>Dry cough___<br>Productive cough___<br>Tiredness / fatigue___<br>Other things you noticed? |
| **Your general medical history** | |
| Have you had any other diagnosed illnesses? | Heart disease___ Liver disease___ Cancers___<br>Diabetes and pre-diabetes___ Anaemia___<br>Hypertension___ Atrial fibrillation___<br>Rheumatoid arthritis___ High cholesterol___<br>Thyroid disease___ Osteoarthritis___ Kidney problems___<br>Other? (please specify) |

| | Medication | Dose | Frequency | Duration |
|---|---|---|---|---|
| What medications do you take for your PF (what dose, how often and for how long have you taken them)? | | | | |
| What other medications* or dietary supplements do you take currently (and what doses)? | Medication | Dose | Frequency | Duration |

| | |
|---|---|
| *(if you take amiodarone or nitrofurantoin, please mention these) | |
| If you have previously been prescribed any regular medications or treatments, what were they? | |
| Have you ever had any surgical operations? | Yes___ No___ |
| Please tell us about the type of operation and the approximate date | |
| Would you like to add any additional comments to add to your answers? | |
| **Your family history** | |
| Have any of these close blood relatives, living or deceased, been diagnosed with a lung disease? | Parent___ grandparent___ brother___ sister____<br>son____ daughter___ uncle____ aunt___<br>first cousin___ nephew___ niece___<br>Other (please specify)_________________________ |
| Were any of the above diagnosed with PF? | Yes_____ No_______ Unsure_______ |
| Were any of the above diagnosed with another ILD (interstitial lung disease)? | Unknown____ CTD-ILD____ RA-ILD____<br>Hypersensitivity Pneumonitis_____ Systemic Scleroderma____ other (please specify)________<br>____________________________________________ |
| Were any of the above diagnosed with another lung disease? | Unknown____ COPD____ Asthma____<br>Lung cancer____ other (please specify)________<br>____________________________________________ |
| Were any of the above diagnosed with rheumatoid arthritis? | Yes_____ No_______ Unsure_______ |
| To the best of your knowledge did either of your parents smoke? | Father: Yes_____ No_______ Unsure_______<br>Mother: Yes_____ No_______ Unsure_______ |

| | |
|---|---|
| How long did they smoke for? | Father: _____years Mother: _____years |
| How many on average did they smoke per day? | Father: packs of cigarettes<br>pipes of tobacco<br>don't know<br>Mother: packs of cigarettes<br>pipes of tobacco<br>don't know |
| To the best of your knowledge, did any of your grandparents smoke? | Paternal grandfather: Yes__ No___ Unsure___<br>Paternal grandmother: Yes__ No___ Unsure___<br><br>Maternal grandfather: Yes__ No___ Unsure___<br>Maternal grandmother: Yes__ No___ Unsure___ |
| How long did they smoke for and roughly how many packs of cigarettes and/or pipes per day? | Paternal grandfather: ___years,___packs___pipes<br>Paternal grandmother: ___years,___packs___pipes<br><br>Maternal grandfather: ___years,___packs___pipes<br>Maternal grandmother: ___years,___packs___pipes |
| Would you like to add any additional comments to add to your answers? | |
| **Your hormone history** | |
| At what age did your periods start? | |
| How many pregnancies have you had? (include miscarriage and terminations) | |
| Do you have any biological children? | Yes___ No___ |
| If yes, how many sons? | |
| If yes, how many daughters? | |
| What was your mother's age at menopause? | |
| Did you have (i) a natural menopause or (ii) a hysterectomy? | |
| Have you had surgery to remove both your ovaries or had chemo/radiotherapy that you were advised could be toxic to the ovaries? | |
| How old were you when first (peri)menopausal symptoms started? (e.g. hot flushes, irregular periods, night sweats…) | |
| How old were you when you had your last period? | |
| Have you ever had any hormone replacement treatment (HRT), including oestrogen, progesterone or a progesterone only coil? | Yes___ No___ |

| | |
|---|---|
| If yes, please complete the following: | |
| Did you have oestrogen only? | Yes___ No___ |
| Did you have oestrogen and progesterone? | Yes___ No___ |
| Did you have progesterone only coil? | Yes___ No___ |
| Did you have additional testosterone medication? | Yes___ No___ |
| Did you have tablets, patches or gel or creams (vaginal or skin)? | |
| At what age did you start HRT? | |
| Are you still taking HRT? | Yes___ No___ |
| If no, at what age did you finish HRT? | |
| What were the worse symptoms for which you sought treatment? | |
| Have you had a hysterectomy?<br>If yes, please complete the following: | Yes___ No___ |
| Approximate date or age of surgery | |
| Symptoms that indicated surgery | |
| Have you had an oophorectomy (ovary removal, single or double)?<br>If yes, please complete the following: | Yes___ No___ |
| Approximate date of surgery | |
| Symptoms that indicated surgery | |
| Would you like to add any additional comments to add to your answers? | |
| **Additional Indications of interest in our research** | |
| Do you have (or are you taking medication for) joint pain/stiffness that lasts longer than half an hour in the mornings? | |
| Roughly how many colds and common infections that you get in a normal year (i.e. that isn't related to COVID isolating)? | |
| Have you noticed any of the following: | |
| (a) Premature hair greying *(defined as before the age of 20 in Europeans, 25 in Asians and age of 30 in Africans)* | Yes___ No___ |
| (b) Nail weakness/splitting/ridging | Yes___ No___ |
| (c) History of skin cancers (particularly face and neck) | |
| (d) Excessive eye tears (e.g. weeping eyes in a gentle breeze) | |
| (e) Slower blood clotting after a cut | |

| | |
|---|---|
| (f) Raynaud's syndrome (ends of fingers go white/numb) | |
| Would you like to add any additional comments to add to your answers? | |
| Is there anything else at all that you'd like to say or would like us to know? | |
| Would you be happy to be contacted for further information about any of your answers? | Yes___ No___ |

**With heartfelt thanks for your contribution to this study!** For further information please use the contact details on the EPIC leaflet.

### Study of Telomeres and Role of Sex Hormones in PF ('STARSHIP') - **Male Patient** Extended Clinical Review Form

The STARSHIP study has been set up to explore the idea that sex hormone therapy might be useful as a treatment option for patients diagnosed with pulmonary fibrosis. Completion of this form will help us to understand this.

**Please answer these questions as fully as you can and are happy to do so.**

| **General questions** | |
|---|---|
| Your unique study identifier: | |
| Designated sex at birth | Male__ Female___ |
| Your age (years) | |
| Today's date (dd/mm/yyyy) | |
| Your postcode *(we use this to calculate things like local pollution level)* | |
| Have you ever smoked? *(please tick or highlight your choice)* | No___ Yes, previously__ Yes, currently__ |
| If yes, in what year did you start smoking? | |
| If yes and you have stopped, in which year? | |
| What did/do you smoke? *(please tick/highlight all that apply)* | Pre-rolled cigarettes____<br>Roll my own cigarettes ___<br>Cigars___<br>Pipe___<br>Vape___<br>Other (please specify)________________ |
| On average, how much did / do you smoke per day (please include grams of tobacco if you know that)? | |
| In which of these bands does your average total household income fall? *(please circle/highlight), (we use this to compare your data with that from our initial study on participants in the UK Biobank).* | 1) <£18000 (2) £18000-£30999<br>(3) £31000-£51999 (4) £52000-£100000<br>(5) >£100000. |
| Do you exercise? *(please tick/highlight)* | Yes___ No___ |
| What type of exercise do you do on a weekly basis? *(please tick/highlight all that apply)* | Yoga___ gentle walking___ brisk walking___<br>cycling___ Pilates___ running ___<br>gardening__ golf__ bowling__ swimming__<br>pulmonary rehabilitation exercises__<br>housework___ other: |
| How long do you spend exercising in an average week? | |
| Do you drink alcohol? | Yes___ No___ |

On average, what alcohol and how many units of each do you drink per week?

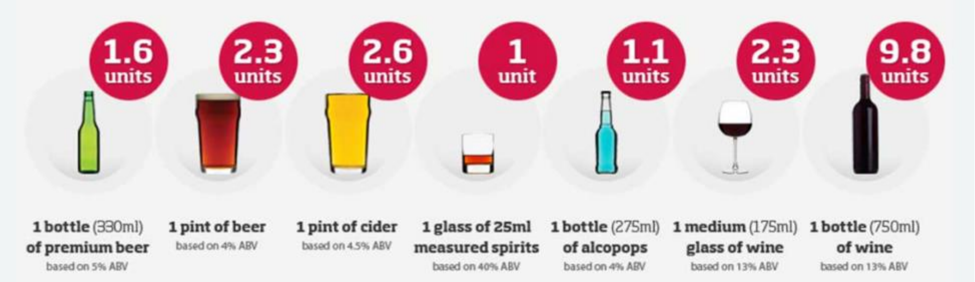

| | |
|---|---|
| Are you retired? | Yes___ No___ |
| What is/was your main occupation(s)? | |
| If you are retired, at what age did you retire? | |
| **Questions about your birth** | |
| To the best of your knowledge, were you born at full term, e.g. between 38-42 weeks of your mother's pregnancy? | Yes___ No___ |
| If no, were you born prematurely (i.e. before 'term' at 37 weeks)? | Yes___ No___ |
| If you were born before full term at how many weeks of your mother's pregnancy were you born? | ________ weeks Not sure |
| What was your approximate birthweight? | ___ lb___oz Don't know____ |
| Do you have any additional comments that you would like to add to any of your answers above? | |

| **Pulmonary Fibrosis** | |
|---|---|
| How old were you when your PF was diagnosed? | |
| Looking back, at what age do you think you had first signs of PF? | |
| What were the early signs and symptoms? | Breathlessness___<br>Dry cough___<br>Productive cough___<br>Tiredness / fatigue___<br>Other things you noticed? |

| | |
|---|---|
| **Your general medical history** | |
| Have you had any other diagnosed illnesses? | Heart disease___ Liver disease___ Cancers___<br>Diabetes and pre-diabetes__ _ Anaemia___<br>Hypertension___ Atrial fibrillation___<br>Rheumatoid arthritis___ High cholesterol___<br>Thyroid disease___ Osteoarthritis___ Kidney problems___<br>Other? (please specify) |
| Have you ever had mumps? | Yes_____ No_______ Unsure_______ |
| Have you had an orchidectomy (testicle removed) or undescended testes? | Yes_____ No_______ Unsure_______ |
| What medications do you take for your PF (what dose, how often and for how long have you taken them)? | Medication Dose Frequency Duration |
| What other medications* or dietary supplements do you take currently (and what doses)?<br><br>*(if you take amiodarone or nitrofurantoin, please mention these) | Medication Dose Frequency Duration |
| If you have previously been prescribed any regular medications or treatments, what were they? | |
| Have you ever had any surgical operations? | Yes___ No___ |

| | |
|---|---|
| Please tell us about the type of operation and the approximate date | |
| Have you had a vasectomy and if so, when? | |
| Would you like to add any additional comments to add to your answers? | |
| **Your family history** | |
| Have any of these close blood relatives, living or deceased, been diagnosed with a lung disease? | Parent___ grandparent___ brother___ sister____<br>son____ daughter___ uncle____ aunt___<br>first cousin___ nephew___ niece___<br>Other (please specify)________________________ |
| Were any of the above diagnosed with PF? | Yes_____ No_______ Unsure_______ |
| Were any of the above diagnosed with another ILD (interstitial lung disease)? | Unknown____ COPD____ Asthma____<br>Lung cancer____ other (please specify)________<br>__________________________________________ |
| Were any of the above diagnosed with rheumatoid arthritis? | Yes_____ No_______ Unsure_______ |
| To the best of your knowledge did either of your parents smoke? | Father: Yes_____ No_______ Unsure_______<br>Mother: Yes_____ No_______ Unsure_______ |
| How long did they smoke for? | Father: _____years Mother: _____years |
| How many on average did they smoke per day? | Father: packs of cigarettes<br>pipes of tobacco<br>don't know<br>Mother: packs of cigarettes<br>pipes of tobacco<br>don't know |
| To the best of your knowledge, did any of your grandparents smoke? | Paternal grandfather: Yes__ No___ Unsure___ -<br>Paternal grandmother: Yes__ No___ Unsure___ |

| | |
|---|---|
| | Maternal grandfather: Yes__ No___ Unsure___<br>Maternal grandmother: Yes__ No___ Unsure___ |
| How long did they smoke for and roughly how many packs of cigarettes and/or pipes per day? | Paternal grandfather: ___years,___packs___pipes<br>Paternal grandmother: ___years,___packs___pipes<br><br>Maternal grandfather: ___years,___packs___pipes<br>Maternal grandmother: ___years,___packs___pipes |
| Would you like to add any additional comments to add to your answers? | |
| **Your hormone history** | |
| At what age did your voice start to change? | |
| Have you had any biological children? | Yes___ No___ |
| If yes, how many sons? | |
| If yes, how many daughters? | |
| Would you like to add any additional comments to add to your answers? | |
| **Additional Indications of interest in our research** | |
| Do you have (or are you taking medication for) joint pain/stiffness that lasts longer than half an hour in the mornings? | |
| Roughly how many colds and common infections that you get in a normal year (i.e. that isn't related to COVID isolating)? | |
| Have you noticed any of the following: | |
| (a) Premature hair greying *(defined as before the age of 20 in Europeans, 25 in Asians and age of 30 in Africans)* | Yes___ No___ |
| (b) Nail weakness/splitting/ridging | Yes___ No___ |
| (c) History of skin cancers (particularly face and neck) | |
| (d) Excessive eye tears (e.g. weeping eyes when it's breezy outside) | |
| (e) Slower blood clotting after a cut | |
| (f) Raynaud's syndrome (ends of fingers go white/numb) | |
| Would you like to add any additional comments to add to your answers? | |

| | |
|---|---|
| Is there anything else at all that you'd like to say or would like us to know? | |
| Would you be happy to be contacted for further information about any of your answers? | Yes___ No___ |

**With heartfelt thanks for your contribution to this study!** For further information please use the contact details on the EPIC leaflet.

### Study of Telomeres and Role of Sex Hormones in PF ('STARSHIP') - **Male Patient** Extended Clinical Review Form

The STARSHIP study has been set up to explore the idea that sex hormone therapy might be useful as a treatment option for patients diagnosed with pulmonary fibrosis. Completion of this form will help us to understand this.

**Please answer these questions as fully as you can and are happy to do so.**

| **General questions** | |
|---|---|
| Your unique study identifier: | |
| Designated sex at birth | Male__M_ Female___ |
| Your age (years) | |
| Today's date (dd/mm/yyyy) | |
| Your postcode *(we use this to calculate things like local pollution level)* | |
| Have you ever smoked? *(please tick or highlight your choice)* | No___ Yes, previously__ Yes, currently__ |
| If yes, in what year did you start smoking? | |
| If yes and you have stopped, in which year? | |
| What did/do you smoke? *(please tick/highlight all that apply)* | Pre-rolled cigarettes____<br>Roll my own cigarettes ___<br>Cigars___<br>Pipe___<br>Vape___<br>Other (please specify)________________ |
| On average, how much did / do you smoke per day (please include grams of tobacco if you know that)? | |
| In which of these bands does your average total household income fall? *(please circle/highlight), (we use this to compare your data with that from our initial study on participants in the UK Biobank).* | 1) <£18000 (2) £18000-£30999<br>(3) £31000-£51999 (4) £52000-£100000<br>(5) >£100000. |
| Do you exercise? *(please tick/highlight)* | Yes___ No___ |
| What type of exercise do you do on a weekly basis? *(please tick/highlight all that apply)* | Yoga___ gentle walking___ brisk walking___<br>cycling___ Pilates___ running ___<br>gardening__ golf__ bowling___swimming__<br>pulmonary rehabilitation exercises__<br>housework___ other: |
| How long do you spend exercising in an average week? | |
| Do you drink alcohol? | Yes___ No___ |

On average, what alcohol and how many units of each do you drink per week?

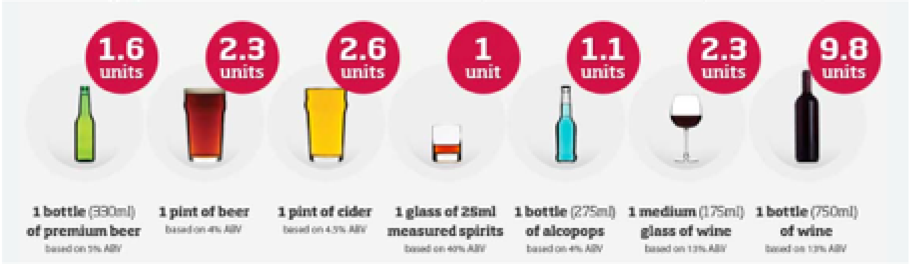

| | |
|---|---|
| Are you retired? | Yes No___ |
| What is/was your main occupation(s)? | |
| If you are retired, at what age did you retire? | |
| **Questions about your birth** | |
| To the best of your knowledge, were you born at full term, e.g. between 38-42 weeks of your mother's pregnancy? | Yes___ No___ |
| If no, were you born prematurely (i.e. before 'term' at 37 weeks)? | Yes___ No___ |
| If you were born before full term at how many weeks of your mother's pregnancy were you born? | _______ weeks Not sure |
| What was your approximate birthweight? | ___ lb___oz Don't know____ |
| Do you have any additional comments that you would like to add to any of your answers above? | |

| **Pulmonary Fibrosis** | |
|---|---|
| How old were you when your PF was diagnosed? | |
| Looking back, at what age do you think you had first signs of PF? | |
| What were the early signs and symptoms? | Breathlessness___<br>Dry cough___<br>Productive cough___<br>Tiredness / fatigue___<br>Other things you noticed? |

| | |
|---|---|
| **Your general medical history** | |
| Have you had any other diagnosed illnesses? | Heart disease ___Liver disease___ Cancers___<br>Diabetes and pre-diabetes___ Anaemia___<br>Hypertension___ Atrial fibrillation___<br>Rheumatoid arthritis_ __ High cholesterol__<br>Thyroid disease___ Osteoarthritis___ Kidney problems___<br>Other? (please specify) |
| Have you ever had mumps? | Yes_____ No_______ Unsure_______ |
| Have you had an orchidectomy (testicle removed) or undescended testes? | Yes_____ No______ Unsure_______ |
| What medications do you take for your PF (what dose, how often and for how long have you taken them)? | Medication Dose Frequency Duration |
| What other medications* or dietary supplements do you take currently (and what doses)?<br><br>*(if you take amiodarone or nitrofurantoin, please mention these) | Medication Dose Frequency Duration |
| If you have previously been prescribed any regular medications or treatments, what were they? | |

| | |
|---|---|
| Have you ever had any surgical operations? | Yes___ No___ |
| Please tell us about the type of operation and the approximate date | |
| Have you had a vasectomy and if so, when? | |
| Would you like to add any additional comments to add to your answers? | |
| **Your family history** | |
| Have any of these close blood relatives, living or deceased, been diagnosed with a lung disease? | Parent___ grandparent___ brother___<br>sister___ son____ daughter___ uncle____<br>aunt___ first cousin___ nephew___ niece___<br>Other (please specify)_________________________<br><br>none |
| Were any of the above diagnosed with PF? | Yes_____ No_______ Unsure_______ |
| Were any of the above diagnosed with another ILD (interstitial lung disease)? | Unknown____ COPD____ Asthma____<br>Lung cancer____ other (please specify)________<br>_____________________________________________ |
| Were any of the above diagnosed with rheumatoid arthritis? | Yes____ _ No_______ Unsure_______ |
| To the best of your knowledge did either of your parents smoke? | Father: Yes_____ No_______ Unsure_______<br>Mother: Yes____ No_______ Unsure_______ |
| How long did they smoke for? | Father: _____years Mother: _____years |
| How many on average did they smoke per day? | Father: packs of cigarettes<br>pipes of tobacco<br>don't know<br>Mother: packs of cigarettes |

| | |
|---|---|
| | pipes of tobacco<br>don't know |
| To the best of your knowledge, did any of your grandparents smoke? | Paternal grandfather: Yes__ No__ Unsure___ -<br>Paternal grandmother: Yes__ No__ Unsure___<br><br>Maternal grandfather: Yes__ No___ Unsure___<br>Maternal grandmother: Yes__ No___ Unsure___ |
| How long did they smoke for and roughly how many packs of cigarettes and/or pipes per day? | Paternal grandfather: ___years, __packs __pipes<br>Paternal grandmother: ___years, __packs __pipes<br><br>Maternal grandfather: ___years, __packs __pipes<br>Maternal grandmother: ___years, __packs __pipes |
| Would you like to add any additional comments to add to your answers? | |
| **Your hormone history** | |
| At what age did your voice start to change? | |
| Have you had any biological children? | |
| If yes, how many sons? | |
| If yes, how many daughters? | |
| Would you like to add any additional comments to add to your answers? | |
| **Additional Indications of interest in our research** | |
| Do you have (or are you taking medication for) joint pain/stiffness that lasts longer than half an hour in the mornings? | |
| Roughly how many colds and common infections that you get in a normal year (i.e. that isn't related to COVID isolating)? | |
| Have you noticed any of the following: | |
| (a) Premature hair greying *(defined as before the age of 20 in Europeans, 25 in Asians and age of 30 in Africans)* | |
| (b) Nail weakness/splitting/ridging | |
| (c) History of skin cancers (particularly face and neck) | |
| (d) Excessive eye tears (e.g. weeping eyes when it's breezy outside) | |

| | |
|---|---|
| (e) Slower blood clotting after a cut | |
| (f) Raynaud's syndrome (ends of fingers go white/numb) | |
| Would you like to add any additional comments to add to your answers? | |
| Is there anything else at all that you'd like to say or would like us to know? | |
| Would you be happy to be contacted for further information about any of your answers? | Yes___ No___ |

**With heartfelt thanks for your contribution to this study!** For further information please use the contact details on the EPIC leaflet.

#### S.2 Safety characteristics; haemoglobin and haematocrit

Haemoglobin (Hb) and haematocrit values were extracted from full blood count results taken on the day of clinic visit to check that treatment with testosterone would be safe in this population (values were unavailable for one male patient). Hb concentrations were within or below the normal range for 97 patients (20/22 female and 77/79 male) and slightly above the normal range for 4 patients (Figure S.3). 67/80 male patients had haemoglobin (Hb) concentration in the normal range (2 above, 13 below). 17/22 female patients had Hb in the normal range (2 above, 3 below) Clinically, a value of 160g/L is used typically as the upper Hb limit for testosterone treatment in males; 90% of the male patients had Hb below this value. It is also the upper limit of normal for females. Mean Hb for all 101 patients was 142 (95%CI:139-146) g/L and mean Hb for 74 controls was the same; 142 (95%CI:140-147) g/L, although the ranges of values were narrower for controls (121-144 g/L for 19 females and 122-172 g/L for 55 males).

Haematocrit measures the ratio of red blood cell volume to total blood volume. The normal range quoted in the UK is 0.37-0.45 for females and 0.40-0.52 for males. 63/80 male patients had haematocrit ratio in the normal range (3 above, 14 below); 15/22 female patients had haematocrit in the normal range (5 above, 2 below). A total of 17/22 (77%) females and 77/79 (97%) males had values below the upper limit. These figures are very similar to those for haemoglobin and there is strong correlation between the two parameters ($\beta$=0.023, $p<10^{-31}$) but those patients with values outside ranges were not identical.

Although lung function (DLCO% or FVC%) was not generally associated with haematocrit ratio after adjusting for age and sex, low DLCO% was associated with having a haematocrit value above the normal range ($\beta$= -15.2 [95%CI: -27.7, -2.6]; p = 0.018). The association for FVC% did not reach significance ($\beta$= -12.5 [95%CI: -27.0, 1.9]; p = 0.087). These results suggest that elevated haematocrit levels above the normal range may be associated with worse lung function amongst ILD patients. It is logical that poor gas transfer might drive haematocrit levels up for improved oxygen uptake and an association between these parameters is reported beyond ILD[3].

The Hb concentration for male patients decreased slightly with age while that for female patients increased slightly. A similar pattern was seen for haematocrit although neither trend reached significance due to sample size (Figures S.2h,k). Linear regression of Hb and haematocrit values in UK biobank data for 170,121 males and 198,247 females of European ethnicity in the age range 40-70 showed similar trends (Figure 3) and this effect has been reported in the literature[4].

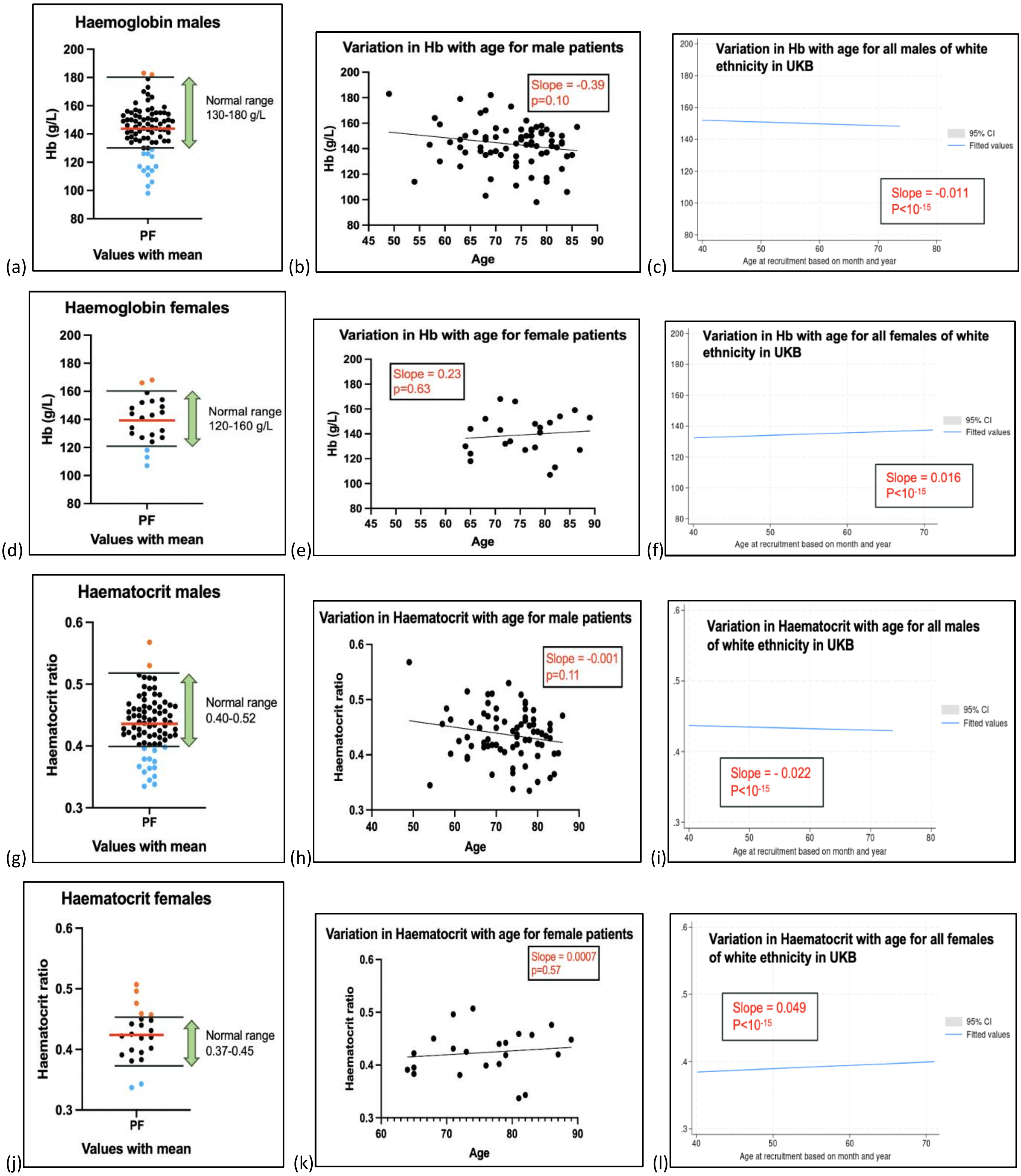

**Figure S.2: Haemoglobin and haematocrit values for male and female patients** (a) Hb for male STARSHIP patients, (b) Hb vs. age for male STARSHIP patients, (c) Hb vs age for UK Biobank males, (d) Hb for female STARSHIP patients, (e) Hb vs. age for female STARSHIP patients, (f) Hb vs age for UK Biobank females, (g) haematocrit for male STARSHIP patients, (h) haematocrit vs age for male STARSHIP patients, (i) haematocrit vs age for UK Biobank males, (j) haematocrit for female STARSHIP patients, (h) haematocrit vs age for female STARSHIP patients, (i) haematocrit vs age for UK Biobank females.

#### S.3 Telomere Length

200/204 samples were analysed using HT-STELA, (n=1 patient sample failed due to a common telomere polymorphism at the 17p telomere; n=3 ASMC samples unable to analyse due to DNA degradation secondary to prolonged storage).

The STARSHIP ASMCs were then combined with pre-existing healthy control samples[1] to provide telomere length age centile curves for 325 healthy controls. Approximately 5% of patients displayed telomeres lengths at or below the first age centile and 20% below the 10th centile (Figures S.3b,c).

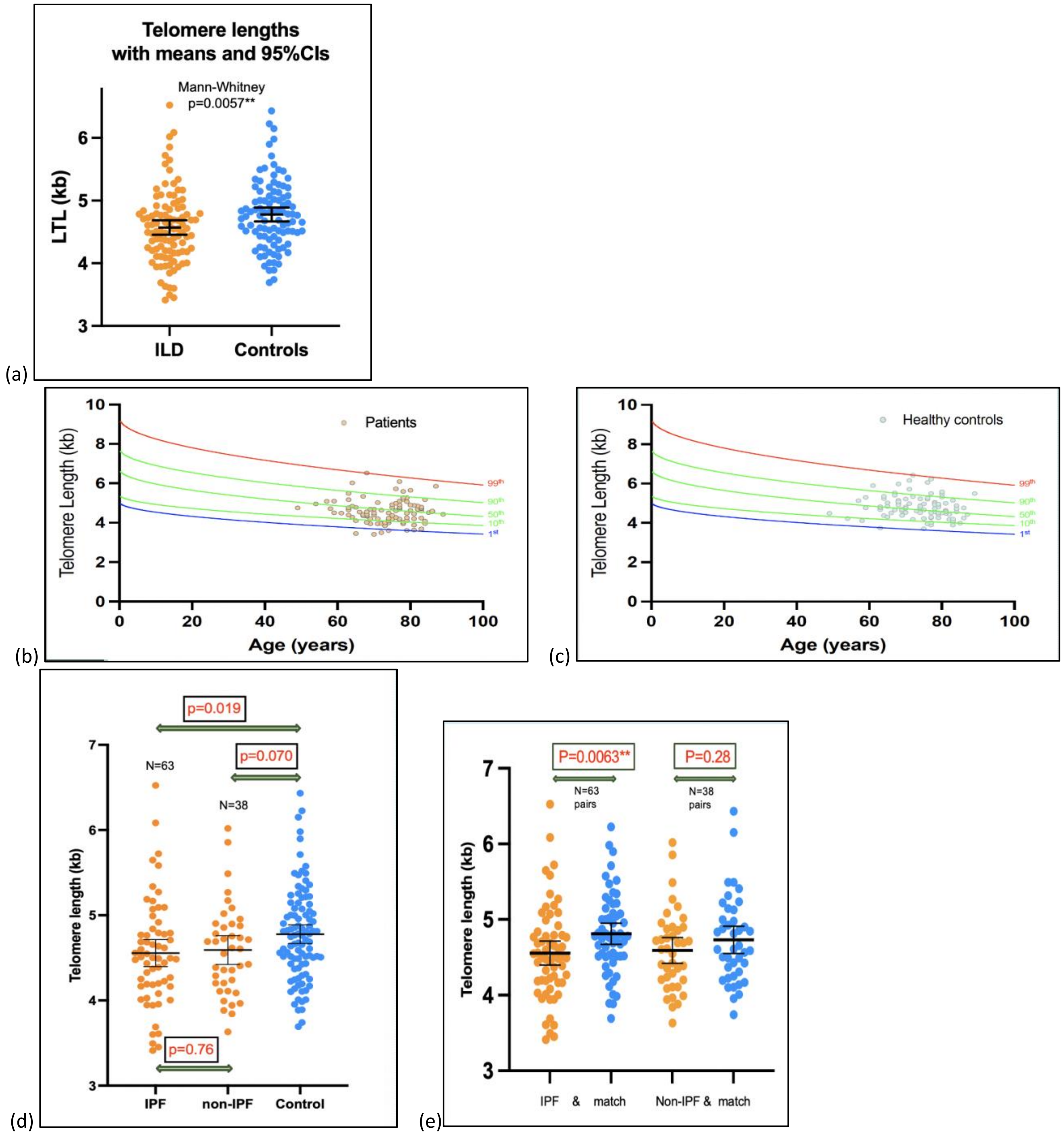

**Figure S.3: Telomere lengths measured using HT-STELA in PF patients and controls**. (a) Leukocyte telomere lengths (LTL) for 101 ILD patients and 99 age and sex matched controls, (b) LTL for PF patients plotted against existing STELA age centiles, (c) LTL for matched controls plotted against existing STELA age centiles, (d) LTL for IPF and non-IPF subgroups of PF compared with all controls, (e) LTL for IPF and non-IPF subgroups compared with their age-matched controls.

Male patients had shorter mean LTL than their ASMCs (p=0.016), but this did not reach significance for the smaller female cohort (Table 3). There was no significant difference in mean LTL between the patient groups with a multi-disciplinary team (MDT) diagnosis of IPF (mean LTL= 4.56kb) and with an MDT diagnosis of non-IPF F-ILD; mean LTL=4.59kb; p=0.76), (Figures 3d,e).

As the HT-STELA technique determines telomere length in bp of DNA, it reveals the telomere length distributions of individual samples. In the absence of telomerase activity, bimodal telomere length distributions are observed that have been attributed to allelic telomere length variation[5]. The presence of telomerase activity creates additional telomere length heterogeneity, the homogenisation of allelic distributions and the loss of allelic bimodality[6]. Bi- or multi-modal telomere length distributions can also arise because of sub-populations of cells with distinct replicative histories and telomere length distributions. Previous analysis of patients with defined mutations in genes required for telomere biology, revealed an increased incidence of bimodal telomere length distributions in affected individuals (15%) compared to normal controls (1.7%), these were attributed to allelic telomere length differentials that were revealed in the context of reduced telomerase activity[1]. Bimodal telomere length distributions were also observed in the STARSHIP cohort with 14/101 (13.9%) ILD cases versus 7/99 (7.1%) controls. Also interestingly, of the ILD cases only 1/22 (4.5%) of the women had a bimodal distribution, compared with 13/79 (16.5%) of the men; none of the female controls had a bimodal distribution. We were not able to formally establish whether this increase in bimodal telomere length distributions was due to allelic telomere length differences in the context of reduced telomerase activity, or from cellular sub-populations with different replicative histories. This phenomenon will be explored in future studies.

**S.4 Impact of prednisolone and smoking and on sex hormones and LTL**

Of the male patients, 12/80 were on oral prednisolone, which is associated with a reduction in circulating testosterone concentrations. Mean total testosterone was lower for those taking prednisolone (13.8 vs 14.6nmol/L), mean bioavailable testosterone was also lower (4.88 vs 5.36nmol/L) but bioavailable testosterone percentage of total testosterone was almost identical (36.4% vs 36.6%) and the free testosterone concentration was slightly higher for those on prednisolone (228pmol/L vs 208pmol/L).

None of the patients interviewed were current smokers. For male patients with PF, total testosterone was slightly lower for those patients who had smoked but this difference was not significant. For the same patient group, SHBG concentration appeared to be raised after adjustments amongst former smokers compared with never smokers, although this did not reach significance $\beta$=8.09 (95%CI: -0.72, 16.9), p=0.072. Lower bioavailable testosterone concentration was associated with former smoking in age and BMI-adjusted regression analysis, $\beta$=-4.75 (95%CI: -8.77, -0.73), p=0.021 for N= 53/72 (73.6%) smokers. Total testosterone was also slightly lower for controls who had smoked but again this difference was not significant. No association for bioavailable testosterone was seen amongst age-matched controls and nor was SHBG concentration positively associated with a history of smoking in this group.

Findings relating to female patients and control groups were not significant. Nor was telomere length associated significantly with a history of smoking in either patients or controls.

For comparison, in the much larger UK Biobank dataset, bioavailable testosterone concentration in 149,096 males was decreased by a history of smoking after adjusting for age and BMI: $\beta$ =-0.065 (95%CI: -0.080, -0.050), $p<10^{-16}$, and SHBG was raised $\beta$ =0.60 (95%CI: 0.45-0.76), $p<10^{-13}$, while the result in 599 male IPF cases was not significant. Again, FAI was unaffected by smoking for 172,769 females and 348 IPF cases in UK Biobank.

The differential effects of smoking on sex hormone concentrations for patients compared with controls were somewhat surprising. If lower bioavailable testosterone increases risk of disease, the reduction of bioavailable testosterone in males might help to explain in part the established association of PF with a history of smoking. This supports our previously reported finding that smoking does not appear to cause IPF directly but does appear to increase exposure to underlying risk[7].

**S.5 Study of albumin concentration in relation to IPF onset**

In order to address the question of whether low albumin is a consequence of or a precursor to disease, we investigated levels amongst incident cases of IPF in UK Biobank data. Mean albumin concentrations at recruitment (in age/sex-adjusted logistic regression) were lower for 916 UK Biobank participants who later developed IPF (mean case albumin [SD] = 43.8 [2.9] g/L, mean control albumin [SD] = 45.2 [2.6] g/L; $p<10^{-36}$ for association between developing IPF and albumin concentration for N=331,471 participants). The time to diagnosis for these cases after albumin measurements at registration was mean (SD) = 4.72 (2.60) years.

The association of hypoalbuminemia with decreased survival has been demonstrated in several conditions, including cancers, end stage renal disease[8] and IPF patients listed for lung transplantation[9]. However, our UK Biobank analysis showed that low albumin concentration may be a pre-clinical feature for people who develop IPF. This could be due to aberrant liver or kidney function, low grade inflammation, malnutrition/suboptimal oral intake, or other symptoms characteristic of a systemic telomere biology disorder[10]. Low albumin predicts mortality in the healthy elderly[11].

##### S.6 Association of LTL with sex hormone levels and lung function with LTL

Telomere length was positively associated with free androgen index in female patients and had a slight negative association for ASMCs (Figure S.6a), with a difference in linear regression slopes of p=0.010. The negative association in controls is explained by a gradual decrease in SHBG and corresponding increase in FAI with age and reflects the expected association of telomere length with age[12]. No association was seen between telomere length and free oestrogen index.

For males a weak association was apparent for telomere length with bioavailable testosterone concentration for patients Figure 6b, but this was not significant either before or after adjusting for age and nor was the difference between slopes of cases and controls (p=0.12).

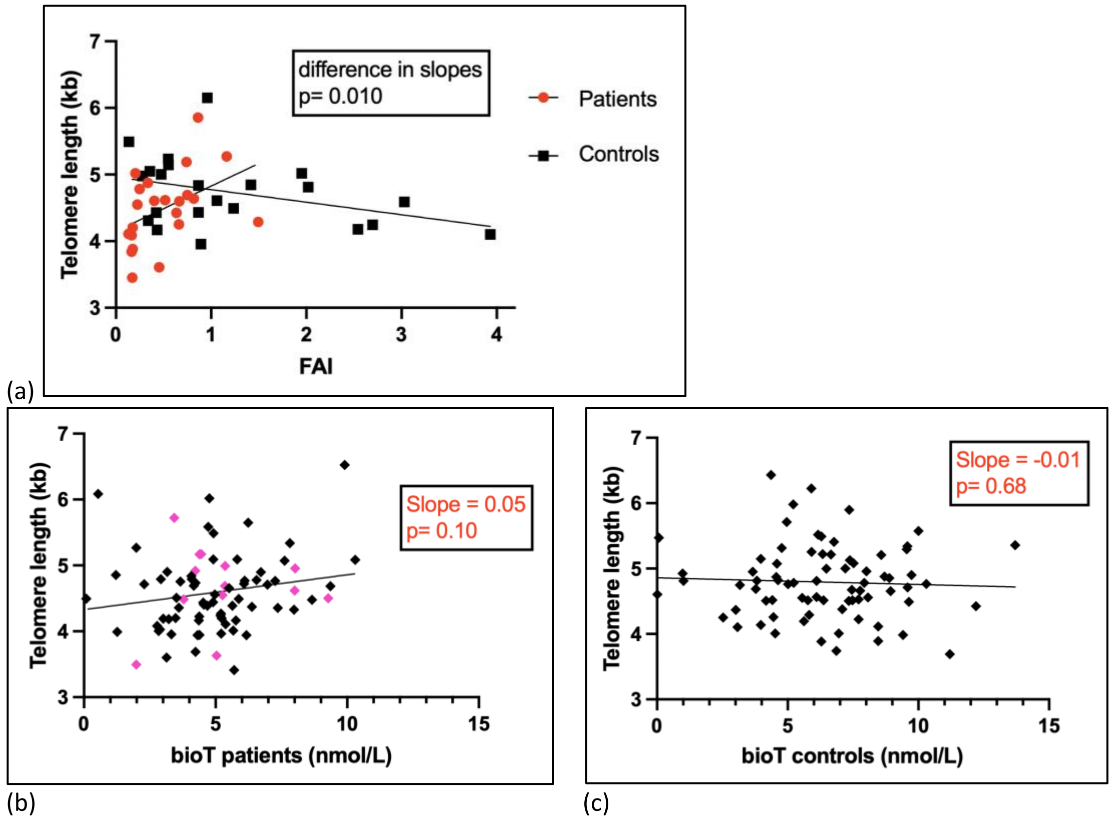

**Figure S.6: Association of leukocyte telomere length with testosterone measures** (a) telomere length association with free androgen index for female PF patients and age-matched controls, (b) telomere length association with bioavailable testosterone concentration in male patients (with pink shading highlighting those with bimodal LTL values, (c) telomere length association with bioavailable testosterone concentration in male age-matched controls.

No association was seen between lung function measures (FVC% predicted and DLCO% predicted), and telomere length in either group of male or female patients. This was the case with both raw data and after adjusting for age (Figure S.7).

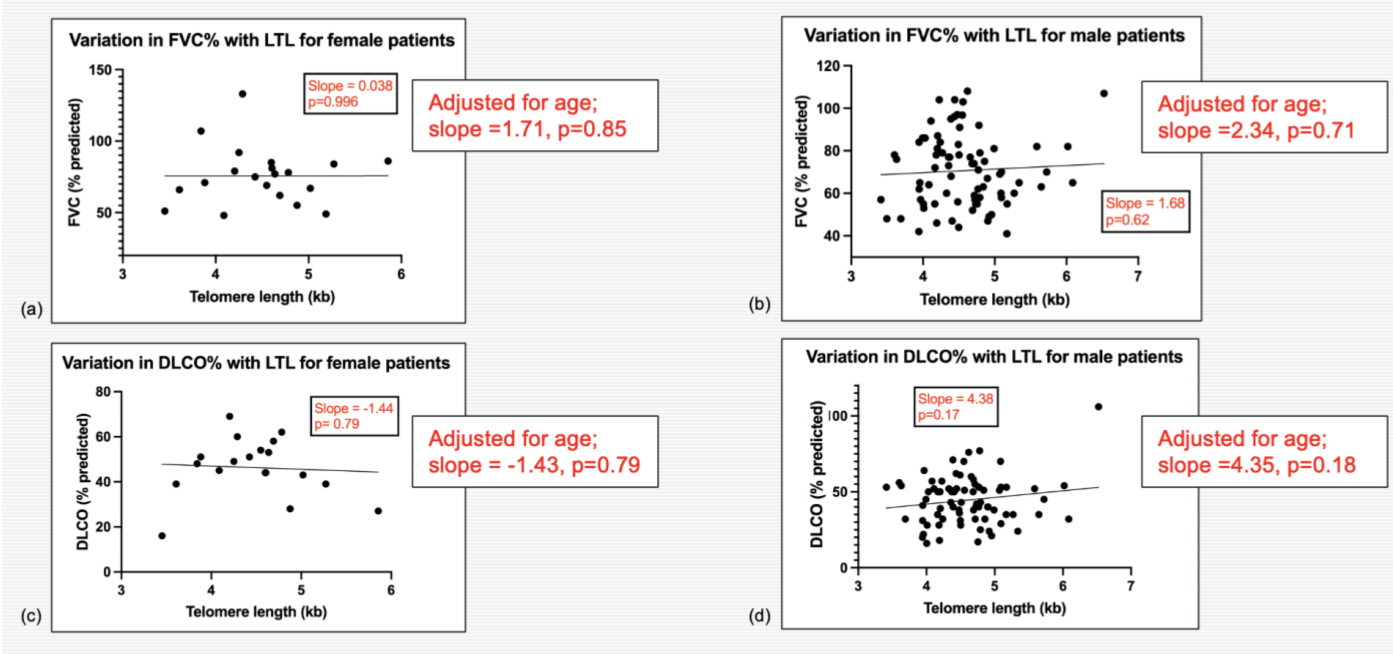

**Figure S.7: Lung function measures, FVC% predicted and DLCO% predicted showed no association with leukocyte telomere length either before or after adjustment for age for females and males in this study.** (a) variation in FVC% predicted with LTL for female patients, (b) variation in FVC% predicted with LTL for male patients, (c) variation in DLCO% predicted with LTL for female patients, (d) variation in DLCO% predicted with LTL for male patients. Regression results adjusted for age are also added in red text for each plot.

#### S.7 Cox proportional hazards modelling for survival and disease progression

**Tables S7a & S7b.** Cox proportional regression analysis of time from study recruitment to death or study census (June 2025) amongst male and female patients with fILD.

| Trait | N | HR | 95% CI | | P value | P test |
|---|---|---|---|---|---|---|
| (i) **Low versus normal free testosterone, adjusted for age, DLCO and FVC** | 78 | **2.23** | **1.05** | **4.75** | **0.038** | 0.11 |
| Age at recruitment | | 0.94 | 0.89 | 0.98 | 0.0033 | |
| DLCO (mL/min/mmHg) | | 0.39 | 0.26 | 0.59 | 7.6x10$^{-6}$ | |
| FVC (L) | | 0.56 | 0.31 | 1.04 | 0.065 | |
| (ii) **Free testosterone (nmol/L), adjusted for age, DLCO and FVC** | 78 | **0.005** | **0.00008** | **0.32** | **0.013** | 0.16 |
| Age at recruitment | | 0.93 | 0.89 | 0.97 | 0.0022 | |
| DLCO (mL/min/mmHg) | | 0.39 | 0.25 | 0.59 | 8.5x10$^{-6}$ | |
| FVC (L) | | 0.62 | 0.33 | 1.18 | 0.14 | |
| (iii) **Low versus normal free testosterone, adjusted for age, DLCO, FVC, IPF, LTL, antifibrotics and immunomodulatory treatments** | 77 | **2.66** | **1.14** | **6.19** | **0.023** | 0.30 |
| Age at recruitment | | 0.92 | 0.87 | 0.97 | 0.001 | |
| DLCO (mL/min/mmHg) | | 0.34 | 0.21 | 0.54 | 7.7x10$^{6}$ | |
| FVC (L) | | 0.48 | 0.23 | 0.98 | 0.044 | |
| IPF diagnosis (score 0 for non-IPF fILD or 1 for IPF) | | 1.22 | 0.58 | 2.60 | 0.60 | |

| | | | | | | |
|---|---|---|---|---|---|---|
| Leukocyte telomere length (Kb) | | 0.81 | 0.44 | 1.48 | 0.49 | |
| Antifibrotic treatment (>1 year) | | 0.60 | 0.22 | 1.68 | 0.33 | |
| Immunomodulatory treatment | | 2.82 | 1.07 | 7.42 | 0.035 | |
| (iv) **Free testosterone (nmol/L), adjusted for age, DLCO, FVC, IPF, LTL, antifibrotics and immunomodulatory treatments** | 72 | **0.0007** | **4.0x10$^{-6}$** | **0.11** | **0.0048** | 0.34 |
| Age at recruitment | | 0.91 | 0.86 | 0.96 | 6.8x10$^{-4}$ | |
| DLCO (mL/min/mmHg) | | 0.32 | 0.20 | 0.52 | 5.9x10$^{-6}$ | |
| FVC (L) | | 0.58 | 0.27 | 1.24 | 0.16 | |
| IPF diagnosis (score 0 for non-IPF fILD or 1 for IPF) | | 1.49 | 0.66 | 3.34 | 0.34 | |
| Leukocyte telomere length (Kb) | | 0.73 | 0.41 | 1.31 | 0.29 | |
| Antifibrotic treatment (>1 year) | | 0.76 | 0.27 | 2.12 | 0.59 | |
| Immunomodulatory treatment | | 3.14 | 1.18 | 8.33 | 0.021 | |

**Table S7a:** Cox proportional hazard analyses of time from recruitment to death or study census amongst males, with respect to (i) Low versus normal free testosterone adjusted for age, absolute DLCO and absolute FVC, (ii) Free testosterone adjusted for age, absolute DLCO and absolute FVC (amongst males and (iii) Low versus normal free testosterone adjusted for age, absolute DLCO, absolute FVC, IPF diagnosis yes/no, leukocyte telomere length, antifibrotic treatment and immunomodulatory treatment, (iv) Free testosterone adjusted for age, absolute DLCO, absolute FVC, IPF diagnosis yes/no, leukocyte telomere length, antifibrotic treatment and immunomodulatory treatment. Testing using Schoenfeld residuals showed no evidence that the proportional hazards assumptions were violated.

| **Trait** | **N** | **HR** | **95% CI** | | **P value** | **P test** |
|---|---|---|---|---|---|---|
| (i) **Below vs above mean FAI, adjusted for age, DLCO and FVC** | 18 | 3.59 | 0.46 | 28.1 | 0.22 | 0.61 |
| Age at recruitment | | 1.02 | 0.85 | 1.22 | 0.84 | |
| DLCO (mL/min/mmHg) | | 0.69 | 0.18 | 2.65 | 0.59 | |
| FVC (L) | | 0.66 | 0.11 | 3.94 | 0.65 | |
| (iii) **Below vs above mean FAI, adjusted for age, DLCO, FVC, IPF, LTL, antifibrotics and immunomodulatory treatments** | 17 | 941 | 0.43 | 20x10$^{5}$ | 0.081 | 0.66 |
| Age at recruitment | | 1.44 | 0.91 | 2.28 | 0.66 | |
| DLCO (mL/min/mmHg) | | 13.4 | 0.14 | 1262 | 0.26 | |
| FVC (L) | | 0.03 | 0.00005 | 13.1 | 0.25 | |
| IPF diagnosis (score 0 for non-IPF fILD or 1 for IPF) | | 4248 | 0.33 | 5.5x10$^{7}$ | 0.084 | |
| Leukocyte telomere length (Kb) | | 7.08 | 0.13 | 392 | 0.34 | |
| Antifibrotic treatment (>1 year) | | 7.24 | 0.28 | 188 | 0.23 | |
| Immunomodulatory treatment | | 5.54 | 0.35 | 87 | 0.22 | |

**Table S7b:** Cox proportional hazard analyses of time from recruitment to death or study census amongst females, with respect to (i) Below versus above mean free androgen index (FAI=0.507%) adjusted for age, absolute DLCO and absolute FVC, (ii) Free androgen index adjusted for age, absolute DLCO, absolute FVC, IPF diagnosis score and leukocyte telomere length. Testing using Schoenfeld residuals showed no evidence that the proportional hazards assumptions were violated.

#### S.8 Excerpts from feedback from two female patients illustrating symptoms

> *"Went on HRT after breaking knee-cap and cracking a couple of ribs, has happened a few times. Started HRT mid 40s and felt a hell of a lot better after, took it for around 15 years, one oestrogen and one progesterone earlier and then just oestrogen later. Would have stayed on it and have asked to go back on it but they wouldn't let me. Stopped probably around 64. Always felt better on it. Now don't feel as bright as used to but it just used to make me feel better. Broke both wrists recently, bones weaker."*

> *"Perimenopausal symptoms in 30s – it was trouble, was on HRT patches, not very nice at all. Was on patches (think both oestrogen and progesterone) for probably 15 years, until had hysterectomy. Had a hysterectomy age 50/51 – was bleeding so heavily and kept bleeding all the time. Stopped HRT around age 55. Treatment was for the hot flushes and night sweats and mood swings. Have type II diabetes since age 65, tablets and insulin twice a day now. Underactive thyroid, been on thyroxine for years, since before age 60."*

**Figure S.8: Hormone related symptoms for two female PF patients.**

#### S.9 Evidence supporting the use of testosterone in treatment of PF

Androgens have been used for several decades to treat more severe telomere biology disorders, showing positive response on growth curves and blood characteristics in children of both sexes[13]. While oestrogen has been shown to have an effect on expression of the telomerase reverse transcriptase (TERT) gene via an oestrogen response element in the TERT promoter region[14], to our knowledge, no independent mechanism for androgen action has been reported and in vitro studies with an androgen receptor agonist made no difference to androgen stimulation of telomerase expression, suggesting that androgen receptors are not involved[14]. Testosterone undergoes downstream conversion to oestrogen via aromatisation, such that oestrogen is also present in low concentrations in males. Aromatisation to oestrogen has been proposed as the method of action for testosterone on telomere length, since tamoxifen (a selective oestrogen receptor modulator) can block the effects of both oestradiol and androgens on telomerase function and such a mechanism is supported by its observed delayed action in vitro compared with oestradiol[14]. The fact that the synthetic androgen danazol cannot be aromatised to oestrogen has been used to argue for an alternative pathway for testosterone activation of telomerase[15]. However, radioactive labelling studies of danazol treatment for endometriosis in women suggest that danazol displaces testosterone from SHBG due to a higher binding affinity and increases the concentration of free testosterone levels compared with pre-treatment levels[16], thus increasing potential for conversion to oestradiol. Danazol also suppresses SHBG levels in plasma[17] so may increase free oestrogen as well. While danazol has been the treatment of choice for telomere-related lung fibrosis patients in several recent trials[18,19], the side-effects in poorly patients have proven unacceptable[20,21].

Current androgen treatment studies for PF and telomere-related diseases have avoided the use of testosterone due to concerns that higher testosterone will increase prostate cancer growth. This stems from a theory that originated with observations in a special population (castrated men) that is not particularly relevant to T therapy in hypogonadal men and has since been discredited[22,23]. Indeed, randomised control trials have reported beneficial effects of testosterone therapy on exercise-induced cardiac ischemia in chronic stable angina, functional exercise capacity, maximum oxygen consumption during exercise (VO2max) and muscle strength in chronic heart failure (CHF), shortening of the Q-T interval, and improvement of some cardiovascular risk factors[24]. A recently published meta-analysis of adverse cardiovascular risk caused by testosterone treatment found no evidence that testosterone increased short-term to medium-term cardiovascular risks in men with hypogonadism[25]. International guidelines are being updated to reflect this finding with the stated proviso that treated men are clinically monitored for adverse effects[26].

There is evidence from meta-analysis of data from 13 studies that that higher testosterone level can significantly decrease the risk of type 2 diabetes in men[27]. In a review of older males with diabetes, a high prevalence of low free testosterone (<225pmol/L) was noted; around 40%, compared with 63% in our STARSHIP study[28]. We have previously reported an association of type II diabetes and short telomeres[29]. In a worldwide audit of testosterone treatment in men with type II diabetes[30], male diabetes patients with low testosterone (N=428 patients entered, mean age 71.4) are receiving testosterone replacement treatment (TRT) as part of normal clinical practice, monitored at 3,6,12 months and then annually after that, in line with TRT guidelines. After 24 months, mean ± SD HbA1c fell by 15.4 ± 8.7 from 71.2 ± 9.3 to 55.8 ± 7.2 mmol/mol (n=101, $p<0.001$). Results from the first four measurement points suggest that TRT progressively improves glycaemic control. The researchers noted that testosterone therapy can also benefit women with type II diabetes.

While oestrogen is the main female sex hormone, testosterone also plays a crucial role in women's health[31]; the mean concentration of circulating testosterone in female serum for the healthy controls in this study was approximately 20x that of oestradiol (609 pmol/L vs 30.5 pmol/L). The 2019 Global Consensus Position Statement on the Use of Testosterone Therapy for Women[32] reports on the scarcity of studies into testosterone action and the effects of testosterone deficiency and replacement in women. The panel highlighted the pressing need for more research into testosterone therapy for women, (particularly for medical conditions other than sexual dysfunction) and the development and licensing of products indicated specifically for women. The British Menopause Society issued guidance for general practitioners on testosterone treatment and monitoring of women in 2020 in response to growing international demand for off-label treatment of hypoactive sexual dysfunction disorder symptomatic women with low testosterone (defined as FAI<1%)[33].

**S.10 Inferences from this study for clinical for telomere length testing in F-ILD**

No association was observed between lung function and telomere length. Mean telomere length in the blood declines slowly with age but changes very little throughout life for each individual; the heritable fraction of leukocyte telomere length is 0.7[34]. The reported association of leukocyte telomere length with survival in PF[35] is more likely an association with strength of genetic predisposition to shorter telomeres than a measure of disease progression and still has importance for clinical management. While the demand for clinical measurement of telomere length as a diagnostic and prognostic biomarker is gaining momentum in the ILD field, this study has highlighted some limitations. The observed overlap between the range of telomere lengths for patients and age-matched controls (Figure 4.3(a)) confirms that single measurements for a patient would have limited diagnostic or prognostic value. For individuals with pathogenic germline telomerase variants, evidence suggests that leukocyte telomere length is short throughout life and if anything, can even get longer with age due to compensatory somatic variations (reported in studies relating to TERT variants[36]) leading to clonal expansion of resultant telomerase-rich hematopoietic stem cells in the bone marrow. The telomere length of patients with defined mutations in telomere biology genes converges with normal in the older age groups, so the ability to discriminate based on telomere length declines as a function of age[1].

This leads to two recommendations for the use of telomere length testing in clinical practice:

1. Telomere length measurement should be carried out if resources allow for patients with non-IPF fibrotic ILD who otherwise may be treated with immunosuppressants, to ensure that their LTL is not below the 10$^{th}$ centile for age, since reports suggest that such treatment could reduce survival[37].

2. If diagnostic testing of familial PF relatives becomes available in future, longitudinal LTL testing should be offered starting at a younger age (say 20-50) when it has greater discriminatory power to highlight variant carrier status for the purposes of pre-clinical treatment and disease prevention and where individual changes over time can be monitored.
